# Using polygenic risk scores for prioritising individuals at greatest need of a CVD risk assessment

**DOI:** 10.1101/2022.10.20.22281120

**Authors:** Ryan Chung, Zhe Xu, Matthew Arnold, Samantha Ip, Hannah Harrison, Jessica Barrett, Lisa Pennells, Lois G. Kim, Emanuele DiAngelantonio, Ellie Paige, Scott C. Ritchie, Michael Inouye, Juliet A. Usher-Smith, Angela M. Wood

**Affiliations:** British Heart Foundation Cardiovascular Epidemiology Unit, Department of Public Health and Primary Care, University of Cambridge, Cambridge, UK; Heart and Lung Research Institute, University of Cambridge, Cambridge, UK; Centre for Cancer Genetic Epidemiology, Department of Public Health and Primary Care, University of Cambridge, Cambridge, UK; Medical Research Council Biostatistics Unit, University of Cambridge, Cambridge, UK; National Institute for Health and Care Research Blood and Transplant Research Unit in Donor Health and Behaviour, University of Cambridge, Cambridge, UK; British Heart Foundation Centre of Research Excellence, University of Cambridge, Cambridge, UK; Health Data Research UK Cambridge, Wellcome Genome Campus and University of Cambridge, Cambridge, UK; Health Data Science Research Centre, Human Technopole, Milan, Italy 20157; National Centre for Epidemiology and Population Health, Australian National University, Canberra, Australia; The George Institute for Global Health, UNSW Sydney, Australia; Cambridge Baker Systems Genomics Initiative, Department of Public Health and Primary Care, University of Cambridge, Cambridge, UK; Cambridge Baker Systems Genomics Initiative, Baker Heart and Diabetes Institute, Melbourne, Victoria, Australia; Primary Care Unit, Department of Public Health and Primary Care, University of Cambridge, Cambridge, UK; Cambridge Centre of Artificial Intelligence in Medicine

## Abstract

**Background:** To provide quantitative evidence of the use of polygenic risk scores (PRS) for systematically identifying individuals for invitation for full formal cardiovascular disease (CVD) risk assessment.

**Methods:** 108,685 participants aged 40-69, with measured biomarkers, linked primary care records and genetic data in UK Biobank were used for model derivation and population health modelling. Prioritisation tools using age, PRS for coronary artery disease and stroke, and conventional risk factors for CVD available within longitudinal primary care records were derived using sex-specific Cox models. Rescaling to account for the healthy cohort effect, we modelled the implications of initiating guideline-recommended statin therapy after prioritising individuals for invitation to a formal CVD risk assessment.

**Results:** 1,838 CVD events were observed over median follow up of 8.2 years. If primary care records were used to prioritise individuals for formal risk assessment using age- and sex-specific thresholds corresponding to 5% false negative rates then we would capture 65% and 43% events amongst men and women respectively. The numbers of men and women needed to be screened to prevent one CVD event (NNS) are 74 and 140 respectively. In contrast, adding PRS to both prioritisation and formal assessments, and selecting thresholds to capture the same number of events resulted in a NNS of 60 for men and 90 for women.

**Conclusion:** The use of PRS together with primary care records to prioritise individuals at highest risk of a CVD event for a formal CVD risk assessment can more efficiently prioritise those who need interventions the most than using primary care records alone. This could lead to better allocation of resources by reducing the number of formal risk assessments in primary care while still preventing the same number CVD events.

## INTRODUCTION

Cardiovascular disease (CVD) remains a major cause of morbidity and mortality worldwide.^1^ Identifying individuals at a high risk of CVD in order to manage and implement interventions to reduce risk of CVD remains an important aim.^2,3^ Prediction tools utilising the risk factor levels of individuals to estimate a 5 or 10 year risk of CVD have been developed to aid clinical decision making and are recommended by healthcare guidelines across the world.^3– 10^ However, recent studies have debated the clinical value and cost effectiveness of national risk assessment programmes.^11–17^ In line with this, recent guidelines have made recommendations to better utilise existing primary care records to improve the stratification of high-risk individuals prior to formal CVD risk assessments^18^. However, few strategies or tools to systematically identify such individuals have been recommended. Proposals have also been recommended to prioritise individuals using CVD-based polygenic risk scores (PRS); such PRS have been shown to be independent of other CVD risk factors, offering improved stratification with high concordance between categories of polygenic risk and future CVD risk across the life course, and to improve discriminatory performance when used to supplement existing CVD risk scores.^19–21^. However, no studies have quantified the impact PRS would have for prioritisation.

Therefore, to investigate the benefits of PRS to systematically prioritise individuals at high risk of CVD, we compare systematically prioritising individuals using a PRS based prioritisation tool against current guidelines recommendations of using a prioritisation tool based on longitudinal primary care records.^22–26^

## METHODS

### UK Biobank data source

Data from UK Biobank (UKB) were used to derive each CVD risk tool (prioritisation and formal assessment) and to model the implications of prioritising individuals for formal assessment. UKB is a prospective cohort study with detailed baseline information, genetic data and linked primary care record data available for 177,359 individuals in England recruited between 2006 and 2010^27^. Genetic data was sequenced using a genome wide array of approximately 826,000 markers with imputation to approximately 96 million markers.^27^ Primary care data was provided from the The Phoenix Partnership, Egton Medical Information Systems and Vision GP system suppliers^28^. Data were linked with secondary care admissions from Hospital Episode Statistics (HES) and mortality records from the Office for National Statistics (ONS). For this study, primary care records were restricted to those measured between the 1^st^ April 2004, the introduction of the Quality and Outcomes Framework (QOF) and UKB baseline survey. To assess the impact of PRS as a prioritisation tool and compare with primary care records, analysis was restricted to individuals with complete genetic data necessary for calculating the PRS, at least one primary care record before baseline survey and without prior CVD or statin initiation. Individuals contributing to the PRS derivation were also excluded.

### CPRD data source

Data from the Clinical Practice Research Datalink (CPRD) were used for estimating the average risk factor values and rescaling of 10-year risks (described in later sections.) The CPRD database is a large UK primary care database containing primary care records^28^ with linked information from HES and mortality records from the ONS. The most recent 5 years available primary care records were extracted for 870,486 individuals who were still alive and without prior CVD on the 1^st^ January 2014 and had no statins throughout follow-up until 31^st^ May 2019, the end of data availability **(Supplementary figure 1)**.

### Outcomes

CVD was defined as the first ever incident of fatal or non-fatal events of coronary heart disease (including angina and myocardial infarction) ischaemic heart disease and stroke (code lists provided in **Supplementary table 1)**, appearing in the linked HES and ONS databases during follow up.

### Risk factors

Two PRS for coronary artery disease (CAD) and stroke, constructed using a meta-score approach and external summary statistics from large genome wide association studies^20,29^, were used as independent variables. Conventional risk factors (as those in the QRISK2 scores^4^) were selected: age, sex, ethnicity, Townsend score, smoking status (current smoker), history of diabetes (type 1 or type 2 or history of diabetes medication), family history of CVD, history of chronic kidney disease (stages 4 and 5), history of atrial fibrillation status, history of blood pressure treatment, history of rheumatoid arthritis, total and high density lipoprotein (HDL) cholesterol, systolic blood pressure (SBP), body mass index (BMI), and age interactions with Townsend score, history of diabetes, family history of CVD, history of atrial fibrillation, history of blood pressure treatment, SBP and BMI.

### Statistical modelling

Sex-specific Cox models were used to derive three different prioritisation tools for estimating 10-year CVD prioritisation risk. First, we derived a prioritisation tool with linear predictors of baseline age, CAD PRS^20^ and stroke PRS^29^. Age interactions were considered but were not statistically significant at the 5% level. Second, we derived a prioritisation tool with predictors utilising longitudinal primary care records. To handle missing values, the tool was derived in two stages: in the first stage, we used sex-specific multivariate mixed effects regression models on longitudinal risk factor measurements for SBP, total and HDL cholesterol and BMI to estimate current risk factor values **(Web appendix 1);** in the second stage, we derived sex-specific Cox models with the estimated current risk factor values for SBP, total and HDL cholesterol and BMI, and the most recent primary care measurements for the remaining QRISK2 risk factors. Third, we derived a prioritisation tool with both PRS and primary care records, using the two-stage approach described above with the addition of linear predictors for the CAD PRS and stroke PRS in the second stage Cox models. For each of these three tools, the model is used to identify individuals crossing a minimum 10-year risk threshold to be invited for a formal assessment.

Sex-specific Cox models were used to derive two formal risk assessment models for predicting 10-year formal assessment CVD risk using risk factor measurements observed at UKB baseline survey. First, we re-derived a model based on QRISK2 predictors and second, we derived a model based on QRISK2 predictors enhanced with the CAD PRS and stroke PRS.

All models were validated using 10-fold cross validation and prognostic ability was quantified using Harrell’s C-index to measure discrimination and the net reclassification improvement (NRI).

### Population Health Modelling

Population health modelling was conducted to compare the population health impact of 1) prioritising using a primary care records-based tool followed by a formal assessment with conventional risk factors, 2) prioritising using a PRS and age-based tool followed by a formal assessment with conventional risk factors and PRS and 3) prioritising using both PRS and primary care records, followed by a formal assessment with conventional risk factors and PRS. **(Figure 1)**. Due to UKB being a cohort of healthier individuals than the UK primary care population, we rescaled all estimated 10-year CVD risks so that the distribution of risks estimated were using age-group- and sex-specific level risk factors obtained from CPRD and the published QRISK2 score to better reflect CVD risk assessment programme in the general population. **(Web appendix 2, supplementary table 2**). Details of the rescaling method has been described elsewhere.^30,31^

**Figure 1:**
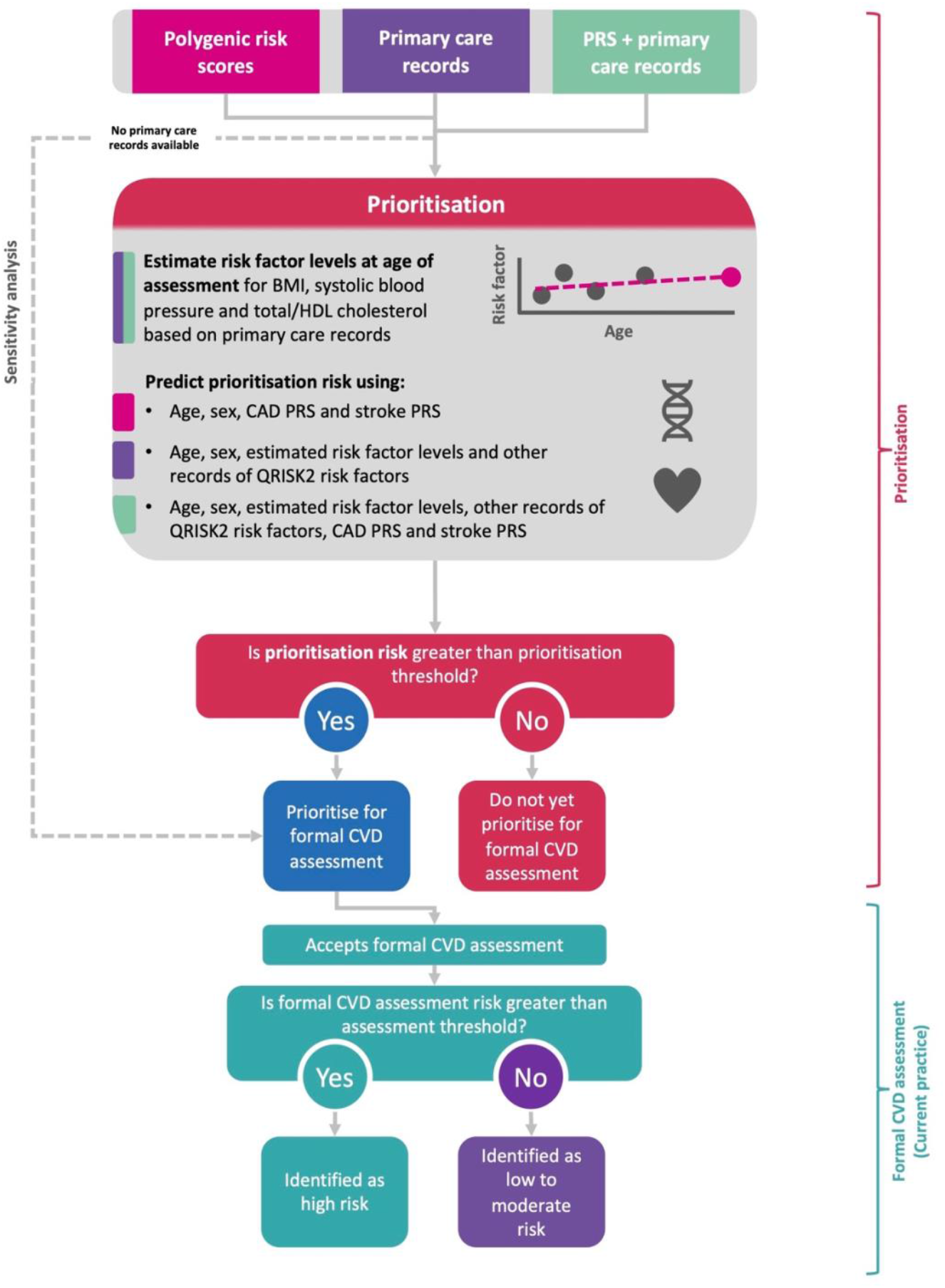
Flow chart of the implementation of a prioritisation tool for formal cardiovascular disease assessments Abbreviations: BMI, body mass index; CAD, coronary artery disease; CVD; cardiovascular disease; HDL, high density lipoprotein; PRS, polygenic risk score.

A hypothetical population of 100,000 individuals (50,000 men and women) from the United Kingdom was created; the population age structure was obtained using data from the ONS in 2015^32^ and the number of expected CVD events was calculated using age-group and sex-specific incidence rates from CPRD **(Supplementary table 3)**. A policy of statin initiation for individuals at ≥10% predicted 10-year formal assessment CVD risk as currently recommended by National Institute for Health and Care Excellence (NICE) guidelines and a 20% reduction in CVD risk were assumed.^33,34^ The population health impact for each of the three prioritisation tools was modelled using age- and sex-specific prioritisation thresholds in two ways. First, we selected prioritisation thresholds to limit the formal CVD risk assessment false negative rate to 5%. Second, we selected prioritisation thresholds for the PRS based prioritisation tools to identify the same number of events that would have been identified if prioritising with primary care records only (with prioritisation thresholds corresponding to 5% false negative rates). We determined the thresholds by varying the false negative rates and chose the rate that best matched the number of events identified.

Summary metrics were estimated for: the number needed to screen (NNS) to prevent one CVD event, the number of CVD events identified and the number needed to invite (NNI) to prevent one CVD event. We assumed 100% statin compliance and a 50% invitation uptake of a formal assessment if inviting all individuals^35,36^. We further assumed an increased invitation uptake of 55% if individuals were prioritised for an invitation to a formal assessment.

In sensitivity analyses, we repeated population-health analyses including all individuals, including those without a primary care record for any one of SBP, HDL, total cholesterol or BMI, where those without a record were all invited for formal assessment **(Supplementary table 3)**. We also repeated analyses assuming that only 50% of individuals treated with statins were compliant with treatment.

Analyses were conducted in R x64 3.6.1 (R Foundation for Statistical Computing, Vienna, Austria). This study follows the RECORD statement (Web Appendix 3)^37^

## RESULTS

### Population characteristics

For our primary analysis, we identified a subset of 108,685 (60%) individuals in UKB with genetic data and a primary care record for at least one of SBP, HDL, total cholesterol and BMI, necessary for the in-person comparison of using either polygenic risk scores or primary care records as a prioritisation method **(Supplementary figure 2)**. All individuals had complete information for the conventional risk factors necessary to calculate a 10-year formal CVD risk at baseline survey.

The mean age at baseline was 56.2 years (SD 8.0) for men and 56.1 years (SD 7.8) for women. During mean of follow-up of 8.2 years, there were 1,838 incident cardiovascular events (**Table 1)**. Compared to the measurements observed at the UKB baseline survey, the measurements recorded in primary care records were lower for SBP and total cholesterol and although similar for current smoking status and history of diabetes, were less concordant for the remaining disease statuses. The mean time between the first available primary care record and baseline survey was 3.8 years (95% CI: 2.3, 4.5) for men and 3.8 years (95% CI: 2.8, 4.7) for women.

**Table 1:** Key characteristics of individuals in UK Biobank baseline survey and linked primary care records

| Characteristic | Men, N = 44,184 (41%) |  | Women, N = 64,501 (59%) |  |
| --- | --- | --- | --- | --- |
| CVD events, N | 1230 |  | 608 |  |
| Follow up duration: years, median (5<sup>th</sup>, 95<sup>th</sup> percentile) | 8.1 (6.1, 10.8) |  | 8.2 (6.8, 10.9) |  |
| Duration between first primary care record and baseline visit: years, median (5<sup>th</sup>, 95<sup>th</sup> percentile) | 3.6 (0.9, 5.5) |  | 3.9 (1.1, 5.6) |  |
|  | Primary care records | Baseline | Primary care records | Baseline |
| Age, mean (SD) <sup>a</sup> | - | 56.2 (8.0) | - | 56.1 (7.8) |
| Coronary artery disease PRS, mean (SD) | - | -1.15 (0.46) | - | -1.13 (0.46) |
| Stroke PRS, mean (SD) | - | 1.55 (0.22) | - | 1.56 (0.23) |
| Ethnicity — White, N (%) <sup>a</sup> | - | 42,283 (95.7%) | - | 61,977 (96.1%) |
| Townsend, mean (SD) <sup>a</sup> | - | -1.5 (3.0) | - | -1.5 (2.9) |
| Systolic blood pressure: mmHg, mean (SD) <sup>b</sup> | 135.3 (7.82) | 141.0 (17.3) | 130.7 (9.62) | 134.9 (19.1) |
| Number of historical records, mean | 3.8 | - | 4.6 | - |
| Total cholesterol: mmol/litre, mean (SD) <sup>b</sup> | 5.48 (0.47) | 5.79 (1.01) | 5.71 (0.50) | 6.03 (1.08) |
| Number of historical records, mean | 2.0 | - | 2.0 | - |
| HDL cholesterol: mmol/litre, mean (SD) <sup>b</sup> | 1.35 (0.19) | 1.30 (0.31) | 1.68 (0.24) | 1.61 (0.37) |
| Number of historical records, mean | 1.8 | - | 1.9 | - |
| BMI: kg/m<sup>2</sup>, mean (SD) <sup>b</sup> | 27.2 (3.1) | 27.5 (4.1) | 26.6 (4.1) | 26.8 (5.0) |
| Number of historical records, mean | 2.0 | - | 2.3 | - |
| Current smoker, N (%) | 4,472 (10.1%) | 5,233 (11.8%) | 4,911 (7.61%) | 5,511 (8.5%) |
| History of diabetes, N (%) | 466 (1.05%) | 630 (1.4%) | 412 (0.64%) | 459 (0.7%) |
| Blood pressure-lowering medication prescriptions, N (%) | 6,396 (14.5%) | 5,529 (12.51%) | 9,737 (15.1%) | 7,643 (11.85%) |
| Family history, N (%) <sup>a</sup> | 1,568 (3.55%) | - | 2,494 (3.87%) | - |
| Chronic kidney disease (4/5), N (%) <sup>a</sup> | 57 (0.13%) | - | 79 (0.12%) | - |
| Rheumatoid arthritis, N (%) | 146 (0.33%) | 381 (0.86%) | 336 (0.52%) | 989 (1.53%) |
| Atrial fibrillation, N (%) | 336 (0.33%) | 123 (0.28%) | 1,749 (2.7%) | 89 (0.14%) |

Abbreviations: BMI, body mass index; CVD, cardiovascular disease; HDL cholesterol, high-density lipoprotein cholesterol; PRS, polygenic risk score; SD, standard deviation.
<sup>a</sup> Risk factor values in both baseline and primary care records if one was missing.
<sup>b</sup> Risk factor values for primary care records estimated using multivariate mixed effects model.

### Model performance and comparison

Hazard ratios (HRs) in the prioritisation tools and formal assessment models, for the same predictors, were similar **(Supplementary table 4-5)**. HRs for the CAD and stroke PRS were higher among men than in women and remained consistent with and without the inclusion of conventional risk factors.

All models had good discriminatory performance with higher performance in women. The C-index for the PRS + age prioritisation tool in men (C-index = 0.663, 95% CI: 0.649, 0.678) was slightly lower than the primary care records prioritisation tool (C-index = 0.684, 95% CI: 0.670, 0.699) **(Table 2)**, however the difference in performance was greater in women. The greatest discriminatory performance was observed in the model using both conventional risk factors with PRS in both men (C-index = 0.716, 95% CI: 0.702, 0.730) and in women (C-index = 0.742, 95% CI: 0.722, 0.762). Using conventional risk factors with PRS also improved the classification of high and low risk individuals compared to using conventional risk factors only in both men (NRI = 0.0262, 95% CI: 0.0072, 0.0458) and in women (NRI = 0.0265, 95% CI: 0.0065, 0.0502) (**Supplementary table 6)**.

**Table 2:** C indices of prioritisation tools and formal CVD risk assessment tools in UK Biobank

| Model | C-index (95% confidence interval) |  |  |
| --- | --- | --- | --- |
|  | All individuals | Men | Women |
| Prioritisation tool |  |  |  |
| Primary care records only | 0.730 (0.719, 0.741) | 0.684 (0.670, 0.699) | 0.734 (0.715, 0.754) |
| PRS + age | 0.663 (0.652, 0.675) | 0.663 (0.649, 0.678) | 0.686 (0.665, 0.707) |
| PRS + primary care records | 0.740 (0.730, 0.751) | 0.704 (0.691, 0.718) | 0.738 (0.718, 0.758) |
| Formal risk assessment tool |  |  |  |
| Conventional risk factors | 0.730 (0.719, 0.740) | 0.700 (0.686, 0.714) | 0.739 (0.720, 0.759) |
| Conventional risk factors +PRS | 0.738 (0.727, 0.749) | 0.716 (0.702, 0.730) | 0.742 (0.722, 0.762) |

Abbreviations: PRS, polygenic risk score.
C indices and 95% confidence intervals from each model for the prediction of 10-year cardiovascular disease by sex and for the combined population in UK Biobank after 10-fold cross validation.

The estimated 10-year risks between the primary care records only prioritisation tool and the formal assessment model using conventional risk factors were highly correlated (correlation coefficient = 0.75 for men and 0.80 for women). In contrast, the estimated 10-year risks between the PRS + age prioritisation tool and the formal assessment model using conventional risk factors and PRS were less highly correlated (correlation coefficients = 0.67 for men and women), and the estimated 10-year risks between the PRS and primary care records based prioritisation tool and the formal assessment model using conventional risk factors with PRS were more highly correlated (correlation coefficients = 0.82 for men and women) **(Table 3)**. Rescaled 10-year risk estimates between all models were similar **(Supplementary figure 3)**.

**Table 3:**
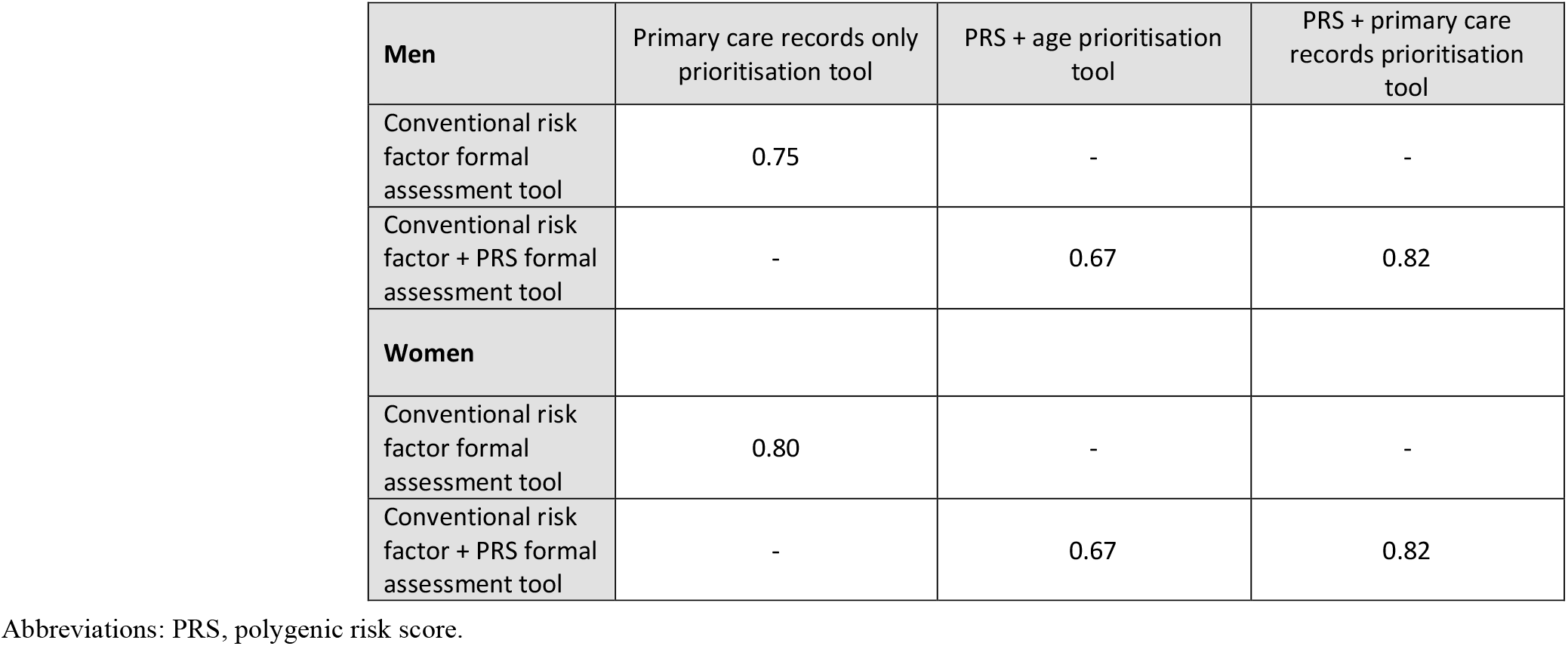
Correlation of predicted 10-year risks between prioritisation tools and formal assessment tools by sex in the derivation dataset

### Population health modelling

In a representative population of 100,000 individuals aged 40 to 69, we estimated that there would be 3,573 men and 1,808 women who would experience a CVD event over the next 10 years. If conventional risk factors were used as a formal risk assessment model on the whole population, then 2,426 (67.9%) of those men and 801 (44.3%) of those women would have been identified as being at high risk, i.e. conventional risk factors ≥ 10% 10-year CVD risk (**Figure 2, supplementary table 7**). Assuming 50% of individuals accepted the invitation for a formal assessment and statin therapy would be initiated on all individuals with a 10-year formal CVD assessment risk greater than 10%, and no other preventive interventions implemented, the NNS to prevent one CVD event in men and women would be 103 (95% CI: 100, 107) and 312 (95% CI: 288, 334), and the NNI to prevent one CVD event in men and women would be 206 (95% CI: 199, 213) and 624 (95% CI: 576, 668) respectively.

**Figure 2:**
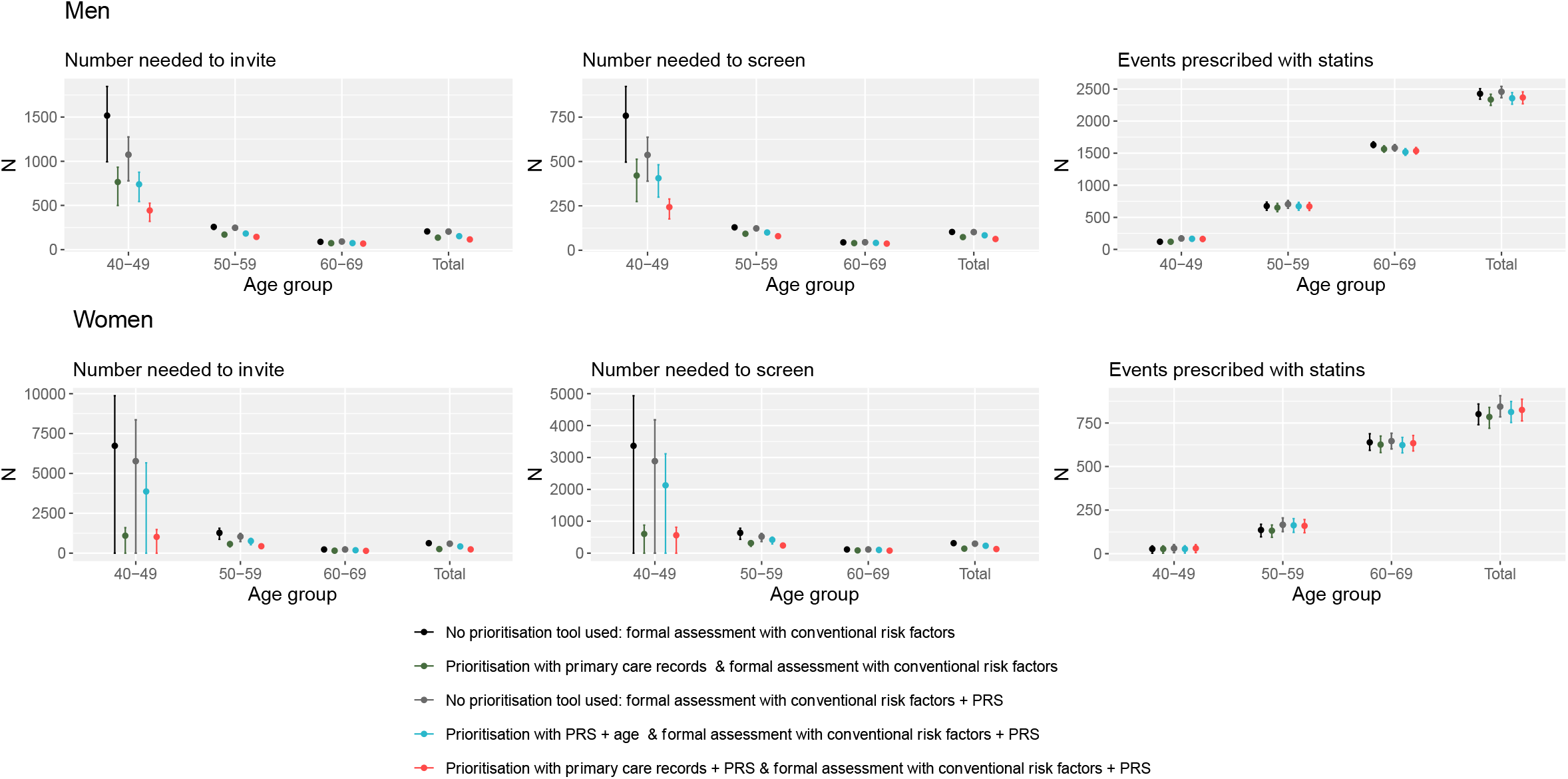
Number needed to invite, number needed to screen and number of events identified after prioritising for a formal CVD assessment, in a hypothetical population of 100,000 individuals in England. Abbreviations: NNS, number needed to screen; NNI, number needed to invite; PRS, polygenic risk score. 95% confidence intervals are represented by vertical lines. Age group and sex specific prioritisation thresholds were defined as the level such that the expected false negative rate is controlled to be 5%. NNI and NNS assumes 100% statin compliance, and half of all individuals invited for formal assessment attend.

If the primary care records-based prioritisation tool, using age- and sex-specific prioritisation thresholds corresponding to 5% false negative rates, was used to prioritise formal assessment (with conventional risk factors) in the population, then 2,335 (65.3%) men and 785 (43.4%) women with CVD events over the next 10 years would be classified at high risk (**Figure 2, supplementary table 7**). The NNS to prevent one event would reduce to 74 (95% CI: 72, 77) in men and 140 (95% CI: 130, 150) in women (28.2% and 55.1% reduction respectively). The NNI to prevent one event would be 135 (95% CI: 130, 141) in men and 255 (95% CI: 235, 274) in women (34.5% and 59.1% reduction respectively)

If conventional risk factors enhanced with PRS was used as a formal risk assessment model on the whole population, then 2,457 (68.8%) of those men and 844 (46.7%) of those women would have been identified as being at high risk **(Figure 2, supplementary table 8)**. The NNS to prevent one CVD event in men and women would be 102 (95% CI: 98, 106) and 296 (95% CI: 273, 315), and the NNI to prevent one CVD event in men and women would be 204 (95% CI: 197, 211) and 592 (95% CI: 545, 631) respectively.

If the PRS + age prioritisation tool, using age- and sex-specific prioritisation thresholds corresponding to 5% false negative rates, was used to prioritise formal risk assessment (with the same conventional risk factors with PRS) in the population, then 78.8% of men and 74.8% of women would be prioritised and, amongst them, 2,356 (65.9%) men and 813 (45.0%) women with CVD events over the next 10 years would be classified at high risk **(Figure 2, supplementary table 8)**. This equates to a 3.7%-4.1% reduction compared to a formal risk assessment using conventional risk factors with PRS on the whole population. However, the NNS to prevent one event would also reduce to 84 (95% CI: 61, 66) in men and 230 (95% CI: 117, 136) in women (17.8% and 22.4% reduction respectively). The NNI to prevent one event would be 152 (95% CI: 146, 158) in men and 418 (95% CI: 384, 447) in women (25.5% and 29.4% reduction respectively).

If the PRS and primary care records-based prioritisation tool, using age- and sex-specific prioritisation thresholds corresponding to 5% false negative rates, was used to prioritise formal assessment (with conventional risk factors with PRS) in the population, then 2,367 (66.3%) men and 825 (45.6%) women would be classified as high risk. **(Figure 2, Supplementary table 9)**. The NNS to prevent one event would reduce to 63 (95% CI: 60, 66) in men and 127 (95% CI: 117, 136) in women (38.2% and 57.1% reduction respectively). The NNI to prevent one event would be 115 (95% CI: 111, 120) in men and 232 (213, 248) in women (43.6% and 60.8% reduction respectively).

Changing the prioritisation thresholds so that the total number of events identified after prioritisation was 63.5% and 43.4% of all events amongst men and women respectively (the total number identified when the primary care records-based prioritisation tool was used with conventional risk factors), prioritising using PRS resulted in a NNS of 82 (95% CI: 79, 85) in men and 223 (95% CI: 205, 240) in women and a NNI of 149 (95% CI: 143, 155) in men and 223 (95% CI: 205, 240) in women. Prioritising using PRS and primary care records resulted in a NNS of 58 (95% CI: 55, 60) in men and 90 (95% CI: 83, 97) in women and a NNI of 105 (95% CI: 101, 110) in men and 163 (95% CI: 151, 176) in women **(Table 4, supplementary figure 4)**. Compared to prioritising using primary care records, the reductions in the NNS and NNI when prioritising using PRS and primary care records, were consistently greater in younger individuals and were statistically significant at the 5% level for all age groups except in women aged 40-49.

**Table 4:** Number needed to invite and screen to prevent one event and number of events identified after prioritisation and formal assessment in a hypothetical population of 100,000 individuals in England, with prioritisation thresholds selected to identify the same number of events if prioritising with primary care records with prioritisation thresholds controlling the false negative rate to 5%.

|  |  | Prioritisation using primary care records followed by conventional risk factors |  |  |  | Prioritisation using PRS + age, followed by conventional risk factors + PRS |  |  |  | Prioritisation using PRS and primary care records, followed by conventional risk factors + PRS |  |  |  |
| --- | --- | --- | --- | --- | --- | --- | --- | --- | --- | --- | --- | --- | --- |
| Age group | Participants | Participants prioritised (%) | NNI (95% CI) | NNS (95% CI) | Number of events identified as high risk (%) | Participants prioritised (%) | NNI (95% CI) | NNS (95% CI) | Number of events identified as high risk (%) | Participants prioritised (%) | NNI (95% CI) | NNS (95% CI) | Number of events identified as high risk (%) |
| **Men** |  |  |  |  |  |  |  |  |  |  |  |  |  |
| 40-49 | 18253 | 10126 (55.5%) | 765 (498.9, 932.8) | 421 (274.4, 513.0) | 120 (24.8%) | 13016 (71.3%) | 727 (530.4, 856.5) | 400 (291.7, 471.1) | 163 (33.6%) | 7208 (39.5%) | 369 (265.8, 438.4) | 203 (146.2, 241.1) | 163 (33.6%) |
| 50-59 | 17391 | 12134 (69.8%) | 170 (150.6, 184.2) | 93 (82.8, 101.3) | 651 (52.5%) | 13100 (75.3%) | 179 (162.6, 193.3) | 98 (89.4, 106.3) | 666 (53.7%) | 9878 (56.8%) | 133 (118.4, 144.4) | 73 (65.1, 79.4) | 654 (52.7%) |
| 60-69 | 14356 | 12517 (87.2%) | 73 (70.4, 74.9) | 40 (38.7, 41.2) | 1564 (84.7%) | 12196 (84.9%) | 74 (70.8, 76.1) | 40 (39.0, 41.9) | 1506 (81.5%) | 11064 (77.1%) | 65 (62.1, 66.9) | 36 (34.2, 36.8) | 1519 (82.3%) |
| Total | 50000 | 34777 (69.6%) | 135 (130.4, 140.5) | 74 (71.7, 77.3) | 2335 (65.3%) | 38313 (76.6%) | 149 (143.0, 154.9) | 82 (78.7, 85.2) | 2335 (65.4%) | 28150 (56.3%) | 105 (100.5, 109.5) | 58 (55.3, 60.2) | 2336 (65.4%) |
| **Women** |  |  |  |  |  |  |  |  |  |  |  |  |  |
| 40-49 | 18107 | 3233 (17.9%) | 1092 (0.0, 1596.1) | 601 (0.0, 877.9) | 27 (10.0%) | 10139 (56.0%) | 3550 (0.0, 5198.6) | 1952 (0.0, 2859.2) | 27 (10.0%) | 1748 (9.7%) | 506 (0.0, 731.1) | 279 (0.0, 402.1) | 31 (11.7%) |
| 50-59 | 17282 | 8329 (48.2%) | 572 (389.4, 701.7) | 314 (214.2, 385.9) | 132 (23.0%) | 12436 (72.0%) | 742 (507.0, 891.7) | 408 (278.8, 490.4) | 156 (27.1%) | 4683 (27.1%) | 306 (198.7, 373.0) | 168 (109.3, 205.2) | 139 (24.1%) |
| 60-69 | 14611 | 10459 (71.6%) | 152 (138.7, 162.1) | 84 (76.3, 89.1) | 626 (65.1%) | 11357 (77.7%) | 178 (163.2, 189.7) | 98 (89.8, 104.3) | 603 (62.7%) | 7714 (52.8%) | 114 (105.2, 121.2) | 63 (57.9, 66.7) | 616 (64.0%) |
| Total | 50000 | 22021 (44.0%) | 255 (235.4, 273.5) | 140 (129.5, 150.4) | 785 (43.4%) | 33932 (67.9%) | 406 (372.3, 436.0) | 223 (204.8, 239.8) | 786 (43.5%) | 14145 (28.3%) | 163 (150.5, 175.5) | 90 (82.8, 96.5) | 786 (43.5%) |

Abbreviations: NNS, number needed to screen; PRS, polygenic risk score.
Age structure of hypothetical population extrapolated from Office for National Statistics, England, United Kingdom 2015. Expected events at 10 years based on extrapolation of incidence rates from CPRD, 2014-2019. Age group and sex specific prioritisation thresholds when prioritising using primary care records were defined as the level such that the expected false negative rate is controlled to be 5%. Age group and sex specific prioritisation thresholds when prioritising using PRS or PRS and primary care records tool chose thresholds that resulted in a similar number of events identified if prioritising using primary care records. Prioritisation thresholds for the PRS prioritisation tool was equivalent to a 5.9% and 6.5% false negative rate for men and women respectively, and for the PRS and primary care records prioritisation tool was equivalent to an 8.1% and 13.0% false negative rate respectively. NNI and NNS assumes 100% statin compliance. NNI assumes a 50% invitation uptake if assessing without using prioritisation tool, and a 55% invitation uptake if assessing with using prioritisation tool.

### Sensitivity Analysis

In sensitivity analyses including all individuals (i.e., including 15,324 individuals without a primary care record for any one of SBP, total cholesterol, HDL cholesterol or BMI) (**Supplementary table 10**), we found comparable results for the PRS-based prioritisation tool and the primary care-based prioritisation tool in men and women. As expected, we observed an increase in the NNS in the prioritisation tools derived with primary care records, especially amongst the lower age group (**Supplementary table 11-13, supplementary figure 5**).

In sensitivity analyses assuming only half of those who accepted the invitation for a formal assessment and were deemed high risk complied with statin uptake, the NNS doubled for all prioritisation tools, with the NNS to prevent one event increasing to 143 in men and 805 in women when using the PRS based prioritisation tool, and 131 in men and 373 in women when using the primary care records-based prioritisation tool (**Supplementary table 14-16, supplementary figure 6)**.

## DISCUSSION

This study has rigorously assessed the impact of using PRS both alone and in combination with traditional risk factors for systematically prioritising individuals for a formal CVD risk assessment, and compared the efficiency and effectiveness against current recommendations of using existing data on CVD risk factors within primary care records. First, we found that adding PRS to both a prioritisation tool and formal CVD risk assessment model improves their correlation, which subsequently leads to higher efficiency and effectiveness of a prioritisation tool, especially amongst younger individuals. Consequently, PRS in combination with primary care records reduces the number of men and women needed to be screened to prevent one CVD event (NNS) by around 20% and 35% respectively, in comparison to using primary care records alone and identifying the same number of events. In contrast, using the PRS alone and in place of primary care records in a prioritisation tool leads to larger NNS. These results support the use of PRS together with primary care records to prioritise individuals at highest risk of a CVD event for a formal CVD risk assessment, which could lead to better allocation of resources by reducing the number of formal risk assessments in primary care.

This study has provided a comparison of prioritisation tools using longitudinal primary care records and/or PRS within the UK population aged between 40 and 69 years who are currently invited for a National Health Service (NHS) Health Check to assess their individual risk of CVD. We have demonstrated the benefits of PRS not only by measuring model discrimination, but also by evaluating the health impact if implemented within this population. Compared with previous studies which have generally focussed on the role of PRS in a formal CVD risk assessment model^20,21,29,38^, our study has uniquely assessed its role in a prioritisation tool, in conjunction with a CVD risk model. We have also shown that if PRS were widely available, the inclusion of PRS in a prioritisation tool could improve the effectiveness of a prioritisation tool especially in younger individuals by reducing the reliance on primary care records.

The benefits in prioritising a subgroup of those individuals at low absolute risk to increase efficiency echoes other studies, which have also shown that selecting a smaller proportion of younger, low-risk individuals can lead to dramatically reduced costs whilst resulting in more Quality Adjusted Life Years (QALYs) gained.^39^ Using a prioritisation tool could also efficiently help reduce the concerning backlog in health checks caused by the COVID-19 pandemic^40^, where the number of people invited to health checks in England declined by 82% between the end of 2019 and 2020,^41–43^ while still preventing nearly the same number of CVD events. Whilst the addition of PRS has the potential to prioritise individuals earlier for a formal CVD assessment, further extensions include using PRS to identify individuals at high risk of other common chronic diseases, including diabetes, dementia and kidney disease.^44–48^

### Strengths and Limitations

Our study has several strengths. This study is the first of our knowledge to directly compare how using different data types for a prioritisation tool can impact on the CVD risk assessment programme in England. This was possible due to the unique data linkage of primary care records along with a baseline survey in UK Biobank. We derived the PRS based prioritisation tools using two current and well documented PRS that have been shown to improve model performance independent of traditional CVD risk factors. We also took advantage of the sporadically observed longitudinal primary care records when deriving the primary care records-based prioritisation tools, by estimating current risk factor values using a multivariate mixed model. Whilst QRISK2, which replaces missing non-recorded values with age, sex and ethnicity-specific population average values, could have been used in our study for the prioritisation tool, we chose to optimise the available longitudinal data in primary care records to reduce possible over-inflation of the information from PRS. Another strength of this study is the use of 10-fold cross validation to correct for over optimism that may exist in our analyses as we derived and conducted the population health modelling in the same individuals. Further, we used rescaling methods to adjust the 10-year risk estimates for all of the models to minimise the healthy selection bias when deriving models in UK Biobank.

However, several potential limitations exist. First, whilst we used primary care records that were no more than six and a half years old before baseline, the mean risk factor levels between primary care records and at the UKB baseline differed within the same individuals which could lead to a different distribution of 10-year risk estimates. This may also weaken the correlations between the prioritisation tool and formal risk assessment models reported. Second, we determined the number of events identified in the population health modelling by calculating the model’s sensitivity in UKB and translating to a hypothetical population; due to the low number of events in UKB, the sensitivity of each model may be limited in accuracy, especially in younger age groups with fewer events. Third, PRS for cardiovascular disease are still under active development and, while we utilise two extensively studied and validated PRS, there are likely more powerful PRS soon to be available^49^. Finally, the age-range of the population health modelling was limited to between 40 and 69 years old due to the use of UK Biobank. This restricts the population health modelling and in particular limits the ability to investigate the early prioritisation capabilities of PRS (which are fixed at conception).

## Conclusion

Population health guidelines in England recommend individuals at higher estimated risk of CVD be prioritised for formal risk assessment. Our results show that incorporating PRS improves the correlation between prioritisation tools and formal CVD risk assessment models. In particular, the use of PRS together with primary care records to prioritise individuals at highest risk of a CVD event for a formal CVD risk assessment has the ability to efficiently prioritise those who need interventions the most, which could lead to better allocation of resources by reducing the number of formal risk assessments in primary care.

## Data Availability

All data files are available from the UK Biobank and CPRD databases.

## DECLARATIONS

## Acknowledgements

CPRD uses data provided by patients and collected by the NHS as part of their care and support. This work was supported by Health Data Research UK, which is funded by the UK Medical Research Council, Engineering and Physical Sciences Research Council, Economic and Social Research Council, Department of Health and Social Care (England), Chief Scientist Office of the Scottish Government Health and Social Care Directorates, Health and Social Care Research and Development Division (Welsh Government), Public Health Agency (Northern Ireland), British Heart Foundation and Wellcome. For the purpose of open access, the author has applied a Creative Commons Attribution (CC BY) licence to any Author Accepted Manuscript version arising from this submission. This work was performed using resources provided by the Cambridge Service for Data Driven Discovery (CSD3) operated by the University of Cambridge Research Computing Service (www.csd3.cam.ac.uk), provided by Dell EMC and Intel using Tier-2 funding from the Engineering and Physical Sciences Research Council (capital grant EP/P020259/1), and DiRAC funding from the Science and Technology Facilities Council (www.dirac.ac.uk).

## Author contributions

R.C., J.A.U. and A.M.W. designed the study. R.C., Z.X., M.A. and A.M.W. were involved in data preparation. R.C., Z.X., M.A., L.P., M.I. and A.M.W were involved with the methodology. R.C., Z.X. and A.M.W. conducted the statistical analysis. R.C. and A.M.W. wrote the first version of the manuscript. All authors reviewed and edited the manuscript.

## Ethics approval

This research has been conducted using the UK Biobank Resource under Application Number 26865. Data from the Clinical Practice Research Datalink (CPRD) were obtained under licence from the UK Medicines and Healthcare products Regulatory Agency (protocol 162RMn2).

## Data availability

All data files are available from the UK Biobank and CPRD databases.

## Transparency

The lead author (RC) affirms that the manuscript is an honest, accurate, and transparent account of the study being reported; that no important aspects of the study have been omitted; and that any discrepancies from the study as planned (and, if relevant, registered) have been explained.

