## Supplementary Tables and Figures for "Using polygenic risk scores for prioritising individuals at greatest need of a CVD risk assessment"

**Supplementary table 1:** Code list used to define cardiovascular disease.

| Endpoint | ICD-10 code |
|---|---|
| Angina pectoris | I20 |
| Myocardial infarction | I21, I22, I23 |
| Coronary disease non- myocardial infarction | I24, I25 |
| Ischemic stroke | I63 |
| Unclassified stroke | I64 |

HES data available covered hospital admissions. Death registries provided data on deaths, with both primary and contributory causes of death coded in ICD-10

**Supplementary table 2:** Age- and sex-specific mean risk factor levels using records from 870,486 individuals in the CPRD database.

| | Men | | | | | | Women | | | | | |
|---|---|---|---|---|---|---|---|---|---|---|---|---|
| Age group | 40-44 | 45-49 | 50-54 | 55-59 | 60-64 | 65-69 | 40-44 | 45-49 | 50-54 | 55-59 | 60-64 | 65-69 |
| Ethnicity — White, (%) | 89.4 | 91.8 | 93.6 | 94.7 | 96 | 97.3 | 88.7 | 90.8 | 92.9 | 94.1 | 95.6 | 96.9 |
| Townsend, mean* | -1.5 | -1.5 | -1.5 | -1.5 | -1.5 | -1.5 | -1.5 | -1.5 | -1.5 | -1.5 | -1.5 | -1.5 |
| Systolic blood pressure — mmHg, mean | 129.5 | 131.2 | 132.8 | 134.4 | 135.8 | 136.5 | 121.0 | 124.3 | 127.5 | 130.0 | 132.6 | 134.8 |
| Total cholesterol — mmol/litre, mean | 5.18 | 5.26 | 5.31 | 5.27 | 5.22 | 5.13 | 4.85 | 5.07 | 5.36 | 5.61 | 5.70 | 5.70 |
| HDL cholesterol — mmol/litre, mean | 1.27 | 1.29 | 1.32 | 1.34 | 1.38 | 1.4 | 1.53 | 1.57 | 1.64 | 1.68 | 1.69 | 1.71 |
| BMI – kg/m2, mean | 28.2 | 28.6 | 28.5 | 28.3 | 27.9 | 27.4 | 27.8 | 28.2 | 28.2 | 28.1 | 27.7 | 27.2 |
| Current smoker, (%) | 49.2 | 47.9 | 45.4 | 40.4 | 33.8 | 27.1 | 42.0 | 43.1 | 43.3 | 39.9 | 32.2 | 26.7 |
| History of diabetes, (%) | 6.0 | 7.0 | 8.3 | 10.2 | 11.6 | 13.4 | 10.1 | 10.5 | 11.0 | 11.3 | 11.7 | 12.6 |
| Blood pressure-lowering medication prescriptions, (%) | 6.9 | 9.9 | 13.7 | 18.7 | 24.2 | 30.5 | 12.3 | 16 | 20.5 | 25.6 | 30.0 | 35.9 |
| Family history, (%) | 6.1 | 7.0 | 7.5 | 7.6 | 7.0 | 6.7 | 7.4 | 0.9 | 9.7 | 10.4 | 10.4 | 9.8 |
| Chronic kidney disease (4/5), (%) | 0.2 | 0.1 | 0.2 | 0.2 | 0.2 | 0.3 | 0.2 | 0.2 | 0.2 | 0.2 | 0.3 | 0.3 |
| Rheumatoid arthritis, (%) | 0.4 | 0.6 | 0.7 | 0.8 | 1.1 | 1.2 | 1.1 | 1.4 | 1.6 | 2.0 | 2.5 | 2.7 |
| Atrial fibrillation (%) | 0.3 | 0.5 | 0.8 | 1.2 | 1.9 | 3.0 | 0.2 | 0.2 | 0.4 | 0.6 | 0.9 | 1.5 |
| Coronary artery disease PRS** | -1.07 | -1.10 | -1.12 | -1.14 | -1.17 | -1.17 | -1.08 | -1.10 | -1.11 | -1.13 | -1.14 | -1.15 |
| Stroke PRS** | 1.59 | 1.58 | 1.57 | 1.56 | 1.154 | 1.54 | 1.59 | 1.58 | 1.57 | 1.56 | 1.55 | 1.55 |

Abbreviations: BMI, body mass index; HDL cholesterol, high-density lipoprotein cholesterol; PRS, polygenic risk score.
*A mean Townsend score of -1.5 from UK Biobank due to insufficient data.
** Mean PRS values from UK Biobank were used due to lack of genetic data in CPRD.

**Supplementary table 3:** Age- and sex-specific crude 10-year cardiovascular disease incidence rates using records from 870,486 individuals in the CPRD database.

| | Men | | | | | | Women | | | | | |
|---|---|---|---|---|---|---|---|---|---|---|---|---|
| Age group | 40-44 | 45-49 | 50-54 | 55-59 | 60-64 | 65-69 | 40-44 | 45-49 | 50-54 | 55-59 | 60-64 | 65-69 |
| Crude incidence rate per 1,000 with at least at least one primary care record of systolic blood pressure, cholesterol, or BMI | 18.2 | 34.4 | 58.1 | 86.6 | 112.3 | 144.6 | 11.5 | 18.0 | 28.6 | 39.0 | 58.1 | 73.3 |
| Crude incidence rate per 1,000 including those without at least one primary care record of systolic blood pressure, cholesterol, or BMI | 17.4 | 33.0 | 56.9 | 83.9 | 109.7 | 142.6 | 11.4 | 17.8 | 28.3 | 39.1 | 58.0 | 72.8 |

Abbreviations: BMI, body mass index.

**Supplementary table 4:** Hazard ratios (95% confidence intervals) for the prioritisation and formal risk assessment tools derived using 44.184 men in UK Biobank.

| **Risk factor** | Primary care records prioritisation tool | PRS + age prioritisation tool | PRS + primary care records prioritisation tool | Conventional risk factor formal assessment tool | Conventional risk factor + PRS formal assessment tool |
|---|---|---|---|---|---|
| Age – per year increase | 1.071 (1.060, 1.083) | 1.070 (1.062, 1.078) | 1.075 (1.063, 1.086) | 1.072 (1.060, 1.083) | 1.075 (1.064, 1.087) |
| Ethnicity – non-White | 0.946 (0.680, 1.316) | NA | 0.464 (0.324, 0.665) | 0.910 (0.654, 1.266) | 0.463 (0.324, 0.663) |
| Townsend | 1.032 (1.012, 1.052) | NA | 1.030 (1.010, 1.050) | 1.029 (1.010, 1.049) | 1.028 (1.008, 1.048) |
| Smoking status – current smoker | 2.060 (1.765, 2.404) | NA | 2.028 (1.737, 2.367) | 1.979 (1.704, 2.298) | 1.957 (1.685, 2.272) |
| Diabetes status - Yes | 1.066 (0.548, 2.074) | NA | 1.040 (0.535, 2.020) | 1.524 (0.968, 2.400) | 1.486 (0.943, 2.341) |
| Chronic kidney disease (stages 4/5) | 2.777 (1.149, 6.709) | NA | 2.691 (1.113, 6.504) | 2.699 (1.118, 6.516) | 2.655 (1.100, 6.412) |
| History of atrial fibrillation - Yes | 0.662 (0.320, 1.368) | NA | 0.651 (0.316, 1.342) | 1.740 (0.614, 4.929) | 1.666 (0.599, 4.631) |
| Anti-hypertensive medication - Yes | 1.087 (0.912, 1.294) | NA | 1.068 (0.897, 1.272) | 1.203 (1.006, 1.439) | 1.169 (0.978, 1.398) |
| Rheumatoid arthritis – Yes | 0.624 (0.201, 1.940) | NA | 0.625 (0.201, 1.942) | 1.614 (1.047, 2.488) | 1.562 (1.013, 2.408) |
| Family history of CVD – Yes | 1.252 (0.941, 1.666) | NA | 1.169 (0.878, 1.556) | 1.254 (0.942, 1.668) | 1.186 (0.891, 1.579) |
| Total cholesterol – per mmol/litre increase | 1.571 (1.386, 1.780) | NA | 1.503 (1.326, 1.705) | 1.303 (1.231, 1.380) | 1.276 (1.205, 1.351) |
| HDL – per mmol/litre increase | 0.373 (0.256, 0.543) | NA | 0.398 (0.274, 0.579) | 0.411 (0.330, 0.511) | 0.424 (0.341, 0.527) |
| Systolic blood pressure – per mmHg increase | 1.025 (1.017, 1.034) | NA | 1.023 (1.014, 1.032) | 1.013 (1.009, 1.016) | 1.012 (1.008, 1.015) |
| BMI – per kg/m2 increase | 1.008 (0.987, 1.030) | NA | 1.007 (0.986, 1.029) | 1.017 (1.002, 1.033) | 1.015 (1.000, 1.031) |
| CAD PRS – per unit increase | NA | 1.769 (1.547, 2.023) | 1.807 (1.575, 2.075) | NA | 1.770 (1.541, 2.032) |
| Stroke PRS – per unit increase | NA | 1.683 (1.291, 2.194) | 1.960 (1.471, 2.612) | NA | 1.868 (1.400, 2.492) |
| Age * BMI | 1.000 (0.997, 1.002) | NA | 1.000 (0.997, 1.002) | 1.000 (0.998, 1.002) | 1.000 (0.998, 1.002) |
| Age * Townsend | 0.998 (0.996, 1.001) | NA | 0.998 (0.995, 1.000) | 0.998 (0.996, 1.001) | 0.998 (0.996, 1.001) |
| Age * Systolic blood pressure | 0.999 (0.998, 1.000) | NA | 0.999 (0.998, 1.000) | 1.000 (0.999, 1.000) | 1.000 (0.999, 1.000) |
| Age * Family history of CVD | 0.991 (0.954, 1.030) | NA | 0.993 (0.955, 1.032) | 0.994 (0.957, 1.032) | 0.996 (0.959, 1.035) |
| Age * Smoking status | 0.984 (0.964, 1.005) | NA | 0.985 (0.965, 1.006) | 0.981 (0.962, 1.000) | 0.982 (0.963, 1.002) |
| Age * Anti-hypertensive medication | 0.994 (0.972, 1.017) | NA | 0.993 (0.971, 1.016) | 0.984 (0.962, 1.008) | 0.983 (0.961, 1.007) |
| Age * Diabetes status | 1.069 (0.988, 1.158) | NA | 1.070 (0.988, 1.158) | 1.016 (0.961, 1.075) | 1.018 (0.962, 1.077) |
| Age * History of atrial fibrillation | 1.060 (0.968, 1.161) | NA | 1.058 (0.966, 1.158) | 1.026 (0.905, 1.164) | 1.024 (0.906, 1.157) |
| Baseline survival estimate at 10 years | 0.9668333 | 0.9670151 | 0.9672866 | 0.9777007 | 0.977394 |

Abbreviations: BMI, body mass index; CAD, coronary artery disease; CVD, cardiovascular disease; HDL cholesterol, high-density lipoprotein cholesterol; PRS, polygenic risk score

**Supplementary table 5:** Hazard ratios (95% confidence intervals) for the prioritisation and formal risk assessment tools derived using 64,501 women in UK Biobank.

| **Rick factor** | Primary care records prioritisation tool | PRS + age prioritisation tool | PRS + primary care records prioritisation tool | Conventional risk factor formal assessment tool | Conventional risk factor + PRS formal assessment tool |
|---|---|---|---|---|---|
| Age – per year increase | 1.065 (1.045, 1.085) | 1.094 (1.081, 1.107) | 1.067 (1.048, 1.087) | 1.076 (1.059, 1.094) | 1.078 (1.061, 1.096) |
| Ethnicity – non-White | 0.936 (0.573, 1.530) | NA | 0.578 (0.338, 0.988) | 0.872 (0.533, 1.426) | 0.562 (0.329, 0.962) |
| Townsend | 1.068 (1.038, 1.100) | NA | 1.067 (1.036, 1.098) | 1.064 (1.034, 1.096) | 1.063 (1.032, 1.094) |
| Smoking status – current smoker | 2.695 (2.134, 3.405) | NA | 2.662 (2.107, 3.362) | 2.528 (2.003, 3.190) | 2.502 (1.983, 3.157) |
| Diabetes status – History or | 1.770 (0.811, 3.861) | NA | 1.771 (0.812, 3.865) | 1.501 (0.670, 3.363) | 1.479 (0.659, 3.319) |
| Chronic kidney disease (stages 4/5) | 0.841 (0.118, 5.993) | NA | 0.795 (0.111, 5.667) | 0.735 (0.102, 5.290) | 0.695 (0.096, 5.021) |
| History of atrial fibrillation - Yes | 0.874 (0.445, 1.717) | NA | 0.881 (0.45, 1.726) | 0.062 (0.000, 19.868) | 0.064 (0.000, 21.315) |
| Anti-hypertensive medication - Yes | 0.995 (0.771, 1.286) | NA | 0.982 (0.76, 1.269) | 1.658 (1.296, 2.123) | 1.622 (1.267, 2.076) |
| Rheumatoid arthritis – Yes | 1.934 (0.917, 4.077) | NA | 1.921 (0.911, 4.05) | 1.479 (0.924, 2.367) | 1.495 (0.934, 2.393) |
| Family history of CVD – Yes | 1.124 (0.734, 1.722) | NA | 1.095 (0.714, 1.677) | 1.097 (0.716, 1.679) | 1.074 (0.701, 1.645) |
| Total cholesterol – per mmol/litre increase | 1.406 (1.181, 1.675) | NA | 1.369 (1.149, 1.631) | 1.227 (1.137, 1.324) | 1.210 (1.121, 1.307) |
| HDL – per mmol/litre increase | 0.365 (0.238, 0.559) | NA | 0.375 (0.245, 0.575) | 0.540 (0.420, 0.694) | 0.547 (0.425, 0.703) |
| Systolic blood pressure – per mmHg increase | 1.038 (1.026, 1.049) | NA | 1.036 (1.025, 1.048) | 1.015 (1.010, 1.020) | 1.014 (1.009, 1.019) |
| BMI – per kg/m2 increase | 0.990 (0.966, 1.014) | NA | 0.990 (0.966, 1.014) | 1.011 (0.994, 1.030) | 1.011 (0.993, 1.029) |
| CAD PRS – per unit increase | NA | 1.424 (1.178, 1.721) | 1.389 (1.144, 1.686) | NA | 1.356 (1.117, 1.647) |
| Stroke PRS – per unit increase | NA | 1.628 (1.113, 2.380) | 1.619 (1.072, 2.445) | NA | 1.562 (1.034, 2.358) |
| Age * BMI | 1.002 (0.999, 1.004) | NA | 1.002 (0.999, 1.004) | 1.001 (0.999, 1.003) | 1.001 (0.999, 1.003) |
| Age * Townsend | 0.998 (0.994, 1.002) | NA | 0.998 (0.994, 1.002) | 0.998 (0.994, 1.001) | 0.998 (0.994, 1.001) |
| Age * Systolic blood pressure | 0.999 (0.998, 1.000) | NA | 0.999 (0.998, 1.000) | 1.000 (0.999, 1.000) | 1.000 (0.999, 1.001) |
| Age * Family history of CVD | 0.974 (0.916, 1.036) | NA | 0.974 (0.916, 1.036) | 0.977 (0.920, 1.037) | 0.977 (0.920, 1.037) |
| Age * Smoking status | 1.009 (0.977, 1.042) | NA | 1.009 (0.977, 1.042) | 1.019 (0.987, 1.051) | 1.018 (0.987, 1.051) |
| Age * Anti-hypertensive medication | 1.005 (0.972, 1.040) | NA | 1.006 (0.972, 1.040) | 0.975 (0.944, 1.007) | 0.975 (0.944, 1.008) |
| Age * Diabetes status | 1.005 (0.904, 1.117) | NA | 1.006 (0.905, 1.118) | 1.030 (0.928, 1.143) | 1.032 (0.930, 1.146) |
| Age * History of atrial fibrillation | 1.066 (0.979, 1.162) | NA | 1.065 (0.978, 1.160) | 1.551 (0.906, 2.653) | 1.540 (0.898, 2.643) |
| Baseline survival estimate at 10 years | 0.9859755 | 0.9896236 | 0.9862068 | 0.9915961 | 0.9915758 |

Abbreviations: BMI, body mass index; CAD, coronary artery disease; CVD, cardiovascular disease; HDL cholesterol, high-density lipoprotein cholesterol; PRS, polygenic risk score.

**Supplementary table 6**: Net reclassification improvement between prioritisation tools and formal risk assessment tools

| Reference | Comparison | Net reclassification improvement (95% confidence interval) | | |
|---|---|---|---|---|
| | | Combined | Men | Women |
| Primary care records only prioritisation tool | PRS + age prioritisation tool | -0.0508 (-0.0787, -0.0299) | -0.0487 (-0.0814, -0.0131) | -0.0870 (-0.1257, -0.0537) |
| Primary care records only prioritisation tool | PRS + primary care records prioritisation tool | 0.0220 (0.0074, 0.0392) | 0.0230 (-0.0005, 0.0481) | 0.0160 (-0.0036, 0.0327) |
| PRS + age prioritisation tool | PRS + primary care records prioritisation tool | 0.0729 (0.0570, 0.0928) | 0.0717 (0.0481, 0.1026) | 0.1030 (0.0765, 0.1346) |
| Conventional risk factor formal assessment tool | Conventional risk factor + PRS formal assessment tool | 0.0237 (0.0078, 0.0409) | 0.0262 (0.0072, 0.0458) | 0.0265 (0.0065, 0.0502) |

Abbreviations: PRS, polygenic risk score.

**Supplementary table 7:** Number needed to invite and screen to prevent one event, and number of events identified when prioritising with primary care records in a hypothetical population of 100,000 individuals in England.

| | | | **No prioritisation tool used: formal assessment with conventional risk factors used for all individuals** | | | **Prioritisation using primary care records, followed by formal assessment with conventional risk factors** | | | |
|---|---|---|---|---|---|---|---|---|---|
| Age group | Participants | Expected number of events in 10 years | NNI (95% CI) | NNS (95% CI) | Number of events identified as high risk (%) | Participants prioritised (%) | NNI (95% CI) | NNS (95% CI) | Number of events identified as high risk (%) |
| **Men** | | | | | | | | | |
| 40-49 | 18253 | 485 | 1517 (993.2, 1846.0) | 758 (496.6, 923.0) | 120 (24.7%) | 10126 (55.5%) | 765 (498.9, 932.8) | 421 (274.4, 513.0) | 120 (24.8%) |
| 50-59 | 17391 | 1240 | 257 (231.9, 280.1) | 129 (116.0, 140.0) | 676 (54.5%) | 12134 (69.8%) | 170 (150.6, 184.2) | 93 (82.8, 101.3) | 651 (52.5%) |
| 60-69 | 14356 | 1847 | 88 (85.5, 90.3) | 44 (42.7, 45.1) | 1629 (88.2%) | 12517 (87.2%) | 73 (70.4, 74.9) | 40 (38.7, 41.2) | 1564 (84.7%) |
| Total | 50000 | 3573 | 206 (199.2, 213.1) | 103 (99.6, 106.6) | 2426 (67.9%) | 34777 (69.6%) | 135 (130.4, 140.5) | 74 (71.7, 77.3) | 2335 (65.3%) |
| **Women** | | | | | | | | | |
| 40-49 | 18107 | 269 | 6731 (0.0, 9871.8) | 3365 (0.0, 4935.9) | 27 (10.0%) | 3233 (17.9%) | 1092 (0.0, 1596.1) | 601 (0.0, 877.9) | 27 (10.0%) |
| 50-59 | 17282 | 577 | 1272 (864.6, 1557.6) | 636 (432.3, 778.8) | 136 (23.6%) | 8329 (48.2%) | 572 (389.4, 701.7) | 314 (214.2, 385.9) | 132 (23.0%) |
| 60-69 | 14611 | 962 | 229 (209.4, 244.0) | 114 (104.7, 122.0) | 639 (66.4%) | 10459 (71.6%) | 152 (138.7, 162.1) | 84 (76.3, 89.1) | 626 (65.1%) |
| Total | 50000 | 1808 | 624 (575.9, 668.0) | 312 (288.0, 334.0) | 801 (44.3%) | 22021 (44.0%) | 255 (235.4, 273.5) | 140 (129.5, 150.4) | 785 (43.4%) |

Abbreviations: NNS, number needed to screen; NNI, number needed to invite.

Age structure of hypothetical population extrapolated from Office for National Statistics, England, United Kingdom 2015. Expected events at 10 years based on extrapolation of incidence rates from CPRD, 2014-2019. Age group and sex specific prioritisation thresholds were defined as the level such that the expected false negative rate was controlled to be 5%. NNI and NNS assumes 100% statin compliance. NNI assumes a 50% invitation uptake if assessing without using prioritisation tool, and a 55% invitation uptake if assessing with using prioritisation tool.

**Supplementary table 8:** Number needed to invite and screen to prevent one event, and number of events identified when prioritising with PRS + age in a hypothetical population of 100,000 individuals in England.

| | | | **No prioritisation tool used: formal assessment with conventional risk factors + PRS used for all individuals** | | | **Prioritisation using PRS + age, followed by formal assessment formal assessment with conventional risk factors + PRS** | | | |
|---|---|---|---|---|---|---|---|---|---|
| Age group | Participants | Expected number of events in 10 years | NNI (95% CI) | NNS (95% CI) | Number of events identified as high risk (%) | Participants prioritised (%) | NNI (95% CI) | NNS (95% CI) | Number of events identified as high risk (%) |
| **Men** | | | | | | | | | |
| 40-49 | 18253 | 485 | 1074 (778.6, 1273.8) | 537 (389.3, 636.9) | 170 (35.1%) | 13525 (74.1%) | 739 (544.2, 876.4) | 406 (299.3, 482.0) | 166 (34.3%) |
| 50-59 | 17391 | 1240 | 247 (224.3, 267.2) | 123 (112.1, 133.6) | 705 (56.9%) | 13456 (77.4%) | 182 (164.4, 197.0) | 100 (90.4, 108.4) | 673 (54.3%) |
| 60-69 | 14356 | 1847 | 91 (87.7, 93.3) | 45 (43.9, 46.7) | 1582 (85.7%) | 12424 (86.5%) | 74 (71.6, 76.9) | 41 (39.4, 42.3) | 1517 (82.1%) |
| Total | 50000 | 3573 | 204 (196.5, 211.0) | 102 (98.2, 105.5) | 2457 (68.8%) | 39405 (78.8%) | 152 (146.2, 157.8) | 84 (80.4, 86.8) | 2356 (65.9%) |
| **Women** | | | | | | | | | |
| 40-49 | 18107 | 269 | 5769 (0.0, 8359.3) | 2885 (0.0, 4179.6) | 31 (11.5%) | 11442 (63.2%) | 3867 (0.0, 5665.8) | 2127 (0.0, 3116.2) | 27 (10.0%) |
| 50-59 | 17282 | 577 | 1038 (727.5, 1236.2) | 519 (363.8, 618.1) | 166 (28.8%) | 13590 (78.6%) | 758 (525.1, 910.4) | 417 (288.8, 500.7) | 163 (28.3%) |
| 60-69 | 14611 | 962 | 226 (209.3, 240.7) | 113 (104.6, 120.3) | 646 (67.2%) | 12357 (84.6%) | 180 (166.7, 191.9) | 99 (91.7, 105.5) | 623 (64.8%) |
| Total | 50000 | 1808 | 592 (545.3, 630.8) | 296 (272.6, 315.4) | 844 (46.7%) | 37389 (74.8%) | 418 (384.3, 446.7) | 230 (211.4, 245.7) | 813 (45.0%) |

Abbreviations: NNS, number needed to screen; NNI, number needed to invite; PRS, polygenic risk score.

Age structure of hypothetical population extrapolated from Office for National Statistics, England, United Kingdom 2015. Expected events at 10 years based on extrapolation of incidence rates from CPRD, 2014-2019. Age group and sex specific prioritisation thresholds were defined as the level such that the expected false negative rate was controlled to be 5%. NNI and NNS assumes 100% statin compliance. NNI assumes a 50% invitation uptake if assessing without using prioritisation tool, and a 55% invitation uptake if assessing with using prioritisation tool.

**Supplementary table 9:** Number needed to invite and screen to prevent one event, and number of events identified when prioritising with PRS and primary care records in a hypothetical population of 100,000 individuals in England.

| | | | No prioritisation tool used: formal assessment with conventional risk factors + PRS used for all individuals | | | Prioritisation using PRS and primary care records, followed by formal assessment with conventional risk factors + PRS | | | |
|---|---|---|---|---|---|---|---|---|---|
| Age group | Participants | Expected number of events in 10 years | NNI (95% CI) | NNS (95% CI) | Number of events identified as high risk (%) | Participants prioritised (%) | NNI (95% CI) | NNS (95% CI) | Number of events identified as high risk (%) |
| **Men** | | | | | | | | | |
| 40-49 | 18253 | 485 | 1074 (778.6, 1273.8) | 537 (389.3, 636.9) | 170 (35.1%) | 7930 (43.4%) | 443 (320.1, 525.5) | 243 (176.0, 289.0) | 163 (33.6%) |
| 50-59 | 17391 | 1240 | 247 (224.3, 267.2) | 123 (112.1, 133.6) | 705 (56.9%) | 10622 (61.1%) | 144 (130.0, 156.1) | 79 (71.5, 85.9) | 670 (54.0%) |
| 60-69 | 14356 | 1847 | 91 (87.7, 93.3) | 45 (43.9, 46.7) | 1582 (85.7%) | 11436 (79.7%) | 68 (65.2, 69.9) | 37 (35.9, 38.4) | 1535 (83.1%) |
| Total | 50000 | 3573 | 204 (196.5, 211.0) | 102 (98.2, 105.5) | 2457 (68.8%) | 29988 (60.0%) | 115 (110.6, 119.7) | 63 (60.8, 65.9) | 2367 (66.3%) |
| **Women** | | | | | | | | | |
| 40-49 | 18107 | 269 | 5769 (0.0, 8359.3) | 2885 (0.0, 4179.6) | 31 (11.5%) | 3513 (19.4%) | 1018 (0.0, 1477.0) | 560 (0.0, 812.3) | 31 (11.7%) |
| 50-59 | 17282 | 577 | 1038 (727.5, 1236.2) | 519 (363.8, 618.1) | 166 (28.8%) | 7616 (44.1%) | 434 (306.9, 519.0) | 239 (168.8, 285.5) | 160 (27.7%) |
| 60-69 | 14611 | 962 | 226 (209.3, 240.7) | 113 (104.6, 120.3) | 646 (67.2%) | 9872 (67.6%) | 142 (130.9, 150.8) | 78 (72.0, 83.0) | 634 (65.9%) |
| Total | 50000 | 1808 | 592 (545.3, 630.8) | 296 (272.6, 315.4) | 844 (46.7%) | 21001 (42.0%) | 232 (213.0, 247.8) | 127 (117.2, 136.3) | 825 (45.6%) |

Abbreviations: NNS, number needed to screen; NNI, number needed to invite; PRS, polygenic risk score.

Age structure of hypothetical population extrapolated from Office for National Statistics, England, United Kingdom 2015. Expected events at 10 years based on extrapolation of incidence rates from CPRD, 2014-2019. Age group and sex specific prioritisation thresholds were defined as the level such that the expected false negative rate was controlled to be 5%. NNI and NNS assumes 100% statin compliance. NNI assumes a 50% invitation uptake if assessing without using prioritisation tool, and a 55% invitation uptake if assessing with using prioritisation tool.

**Supplementary table 10:** Summary of number of individuals without primary care records in UK Biobank

| Sex | Age group | Individuals without at least one CVD risk factor in primary care record N (%) |
|---|---|---|
| Men | 40-49 | 3851 (25.0%) |
| | 50-59 | 2736 (14.8%) |
| | 60-69 | 1734 (9.3%) |
| Women | 40-49 | 2714 (14.0%) |
| | 50-59 | 2415 (9.2%) |
| | 60-69 | 1874 (7.2%) |

Abbreviations: CVD, cardiovascular disease.

Prioritisation with primary care records requires at least one CVD risk factor of: systolic blood pressure, total cholesterol, HDL cholesterol and/or BMI.

**Supplementary table 11:** Number needed to invite and screen to prevent one event and number of events identified when prioritising using primary care records, including all individuals without a primary care record for any one of SBP, HDL, total cholesterol or BMI, in a hypothetical population of 100,000 individuals in England.

| | | | **No prioritisation tool used: formal assessment with conventional risk factors used for all individuals** | | | **Prioritisation using primary care records, followed by formal assessment with conventional risk factors** | | | |
|---|---|---|---|---|---|---|---|---|---|
| Age group | Participants | Expected number of events in 10 years | NNI (95% CI) | NNS (95% CI) | Number of events identified as high risk (%) | Participants prioritised (%) | NNI (95% CI) | NNS (95% CI) | Number of events identified as high risk (%) |
| **Men** | | | | | | | | | |
| 40-49 | 18253 | 465 | 1521 (1063.2, 1855.4) | 761 (531.6, 927.7) | 120 (25.8%) | 12154 (66.6%) | 921 (650.9, 1122.5) | 506 (358.0, 617.4) | 120 (25.8%) |
| 50-59 | 17391 | 1207 | 268 (243.9, 288.6) | 134 (121.9, 144.3) | 650 (53.9%) | 12914 (74.3%) | 187 (170.0, 202.3) | 103 (93.5, 111.2) | 628 (52.0%) |
| 60-69 | 14356 | 1814 | 89 (86.8, 91.2) | 45 (43.4, 45.6) | 1611 (88.8%) | 12688 (88.4%) | 74 (72.1, 76.2) | 41 (39.6, 41.9) | 1551 (85.5%) |
| Total | 50000 | 3486 | 210 (203.6, 216.9) | 105 (101.8, 108.5) | 2381 (68.3%) | 37756 (75.5%) | 149 (144.3, 154.4) | 82 (79.3, 84.9) | 2300 (66.0%) |
| **Women** | | | | | | | | | |
| 40-49 | 18107 | 267 | 7134 (0.0, 10223.4) | 3567 (0.0, 5111.7) | 25 (9.4%) | 5316 (29.4%) | 1904 (0.0, 2729.6) | 1047 (0.0, 1501.3) | 25 (9.5%) |
| 50-59 | 17282 | 575 | 1263 (751.0, 1511.9) | 632 (375.5, 756.0) | 137 (23.8%) | 9153 (53.0%) | 622 (365.9, 750.8) | 342 (201.3, 413.0) | 134 (23.3%) |
| 60-69 | 14611 | 957 | 232 (217.0, 247.4) | 116 (108.5, 123.7) | 629 (65.7%) | 10760 (73.6%) | 158 (147.9, 169.4) | 87 (81.3, 93.2) | 617 (64.5%) |
| Total | 50000 | 1799 | 632 (583.8, 675.3) | 316 (291.9, 337.6) | 792 (44.0%) | 25229 (50.5%) | 295 (271.5, 316.8) | 162 (149.3, 174.3) | 776 (43.2%) |

Abbreviations: HDL, high-density lipoprotein; NNI, number needed to invite; NNS, number needed to screen; SBP, systolic blood pressure.

Age structure of hypothetical population extrapolated from Office for National Statistics, England, United Kingdom 2015. Expected events at 10 years based on extrapolation of incidence rates from CPRD, 2014-2019. Age group and sex specific prioritisation thresholds were defined as the level such that the expected false negative rate was controlled to be 5%. NNI and NNS assumes 100% statin compliance. NNI assumes a 50% invitation uptake if assessing without using prioritisation tool, and a 55% invitation uptake if assessing with using prioritisation tool.

**Supplementary table 12:** Number need to invite and screen to prevent one event, and number of events identified when prioritising using PRS + age, including all individuals without a primary care record for any one of SBP, HDL, total cholesterol or BMI, in a hypothetical population of 100,000 individuals in England.

| | | | **No prioritisation tool used: formal assessment with conventional risk factors + PRS used for all individuals** | | | **Prioritisation using PRS + age, followed by formal assessment with conventional risk factors + PRS** | | | |
|---|---|---|---|---|---|---|---|---|---|
| Age group | Participants | Expected number of events in 10 years | NNI (95% CI) | NNS (95% CI) | Number of events identified as high risk (%) | Participants prioritised (%) | NNI (95% CI) | NNS (95% CI) | Number of events identified as high risk (%) |
| **Men** | | | | | | | | | |
| 40-49 | 18253 | 465 | 1141 (858.8, 1328.4) | 570 (429.4, 664.2) | 160 (34.4%) | 13287 (72.8%) | 767 (575.3, 901.4) | 422 (316.4, 495.8) | 157 (33.9%) |
| 50-59 | 17391 | 1207 | 253 (230.3, 271.7) | 126 (115.1, 135.9) | 688 (57.0%) | 13338 (76.7%) | 184 (168.0, 198.7) | 101 (92.4, 109.3) | 658 (54.5%) |
| 60-69 | 14356 | 1814 | 92 (89.0, 94.4) | 46 (44.5, 47.2) | 1563 (86.2%) | 12393 (86.3%) | 75 (72.7, 78.0) | 41 (40.0, 42.9) | 1495 (82.4%) |
| Total | 50000 | 3486 | 207 (200.1, 213.9) | 104 (100.0, 107.0) | 2411 (69.2%) | 39018 (78.0%) | 154 (148.0, 158.3) | 84 (81.4, 87.1) | 2310 (66.3%) |
| **Women** | | | | | | | | | |
| 40-49 | 18107 | 267 | 6115 (0.0, 8605.7) | 3057 (0.0, 4302.9) | 30 (11.2%) | 11224 (62.0%) | 4020 (0.0, 5766.0) | 2211 (0.0, 3171.3) | 25 (9.5%) |
| 50-59 | 17282 | 575 | 1049 (745.3, 1231.3) | 524 (372.7, 615.6) | 165 (28.7%) | 13545 (78.4%) | 776 (540.8, 918.5) | 427 (297.5, 505.2) | 159 (27.6%) |
| 60-69 | 14611 | 957 | 230 (215.3, 244.8) | 115 (107.6, 122.4) | 634 (66.2%) | 12341 (84.5%) | 184 (170.6, 196.5) | 101 (93.8, 108.1) | 610 (63.8%) |
| Total | 50000 | 1799 | 603 (555.0, 646.9) | 302 (277.5, 323.5) | 829 (46.1%) | 37110 (74.2%) | 425 (391.6, 457.0) | 234 (215.4, 251.4) | 794 (44.1%) |

Abbreviations: HDL, high-density lipoprotein; NNI, number needed to invite; NNS, number needed to screen; PRS, polygenic risk score; SBP, systolic blood pressure.

Age structure of hypothetical population extrapolated from Office for National Statistics, England, United Kingdom 2015. Expected events at 10 years based on extrapolation of incidence rates from CPRD, 2014-2019. Age group and sex specific prioritisation thresholds were defined as the level such that the expected false negative rate was controlled to be 5%. NNI and NNS assumes 100% statin compliance. NNI assumes a 50% invitation uptake if assessing without using prioritisation tool, and a 55% invitation uptake if assessing with using prioritisation tool.

**Supplementary table 13:** Number need to invite and screen to prevent one event, and number of events identified when prioritising using PRS and primary care records, including all individuals without a primary care record for any one of SBP, HDL, total cholesterol or BMI, in a hypothetical population of 100,000 individuals in England.

| | | | **No prioritisation tool used: formal assessment with conventional risk factors + PRS used for all individuals** | | | **Prioritisation using PRS and primary care records, followed by formal assessment with conventional risk factors + PRS** | | | |
|---|---|---|---|---|---|---|---|---|---|
| Age group | Participants | Expected number of events in 10 years | NNI (95% CI) | NNS (95% CI) | Number of events identified as high risk (%) | Participants prioritised (%) | NNI (95% CI) | NNS (95% CI) | Number of events identified as high risk (%) |
| **Men** | | | | | | | | | |
| 40-49 | 18253 | 465 | 1141 (858.8, 1328.4) | 570 (429.4, 664.2) | 160 (34.4%) | 10505 (57.6%) | 616 (465.7, 725.7) | 339 (256.2, 399.1) | 155 (33.3%) |
| 50-59 | 17391 | 1207 | 253 (230.3, 271.7) | 126 (115.1, 135.9) | 688 (57.0%) | 11626 (66.9%) | 161 (145.2, 173.4) | 88 (79.9, 95.3) | 658 (54.5%) |
| 60-69 | 14356 | 1814 | 92 (89.0, 94.4) | 46 (44.5, 47.2) | 1563 (86.2%) | 11708 (81.6%) | 70 (67.5, 72.2) | 38 (37.1, 39.7) | 1521 (83.8%) |
| Total | 50000 | 3486 | 207 (200.1, 213.9) | 104 (100.0, 107.0) | 2411 (69.2%) | 33840 (67.7%) | 132 (127.2, 136.6) | 72 (70.0, 75.1) | 2334 (66.9%) |
| **Women** | | | | | | | | | |
| 40-49 | 18107 | 267 | 6115 (0.0, 8605.7) | 3057 (0.0, 4302.9) | 30 (11.2%) | 5557 (30.7%) | 1706 (0.0, 2398.1) | 938 (0.0, 1318.9) | 30 (11.1%) |
| 50-59 | 17282 | 575 | 1049 (745.3, 1231.3) | 524 (372.7, 615.6) | 165 (28.7%) | 8506 (49.2%) | 488 (337.2, 571.9) | 268 (185.5, 314.6) | 159 (27.6%) |
| 60-69 | 14611 | 957 | 230 (215.3, 244.8) | 115 (107.6, 122.4) | 634 (66.2%) | 10215 (69.9%) | 149 (139.3, 158.8) | 82 (76.6, 87.3) | 622 (65.0%) |
| Total | 50000 | 1799 | 603 (555.0, 646.9) | 302 (277.5, 323.5) | 829 (46.1%) | 24278 (48.6%) | 272 (249.9, 291.3) | 150 (137.4, 160.2) | 810 (45.0%) |

Abbreviations: HDL, high-density lipoprotein; NNI, number needed to invite; NNS, number needed to screen; PRS, polygenic risk score; SBP, systolic blood pressure.

Age structure of hypothetical population extrapolated from Office for National Statistics, England, United Kingdom 2015. Expected events at 10 years based on extrapolation of incidence rates from CPRD, 2014-2019. Age group and sex specific prioritisation thresholds were defined as the level such that the expected false negative rate was controlled to be 5%. NNI and NNS assumes 100% statin compliance. NNI assumes a 50% invitation uptake if assessing without using prioritisation tool, and a 55% invitation uptake if assessing with using prioritisation tool.

**Supplementary table 14:** Number need to invite and screen to prevent one event and number of events identified when prioritising using primary care records, assuming statin compliance of 50%, in a hypothetical population of 100,000 individuals in England.

| | | | **No prioritisation tool used: formal assessment with conventional risk factors used for all individuals** | | | **Prioritisation using primary care records, followed by formal assessment with conventional risk factors** | | | |
|---|---|---|---|---|---|---|---|---|---|
| Age group | Participants | Expected number of events in 10 years | NNI (95% CI) | NNS (95% CI) | Number of events identified as high risk (%) | Participants prioritised (%) | NNI (95% CI) | NNS (95% CI) | Number of events identified as high risk (%) |
| **Men** | | | | | | | | | |
| 40-49 | 18253 | 485 | 3033 (1986.4, 3692.0) | 1517 (993.2, 1846.0) | 120 (24.7%) | 10126 (55.5%) | 1530 (997.9, 1865.6) | 841 (548.9, 1026.1) | 120 (24.8%) |
| 50-59 | 17391 | 1240 | 515 (463.8, 560.2) | 257 (231.9, 280.1) | 676 (54.5%) | 12134 (69.8%) | 339 (301.2, 368.3) | 187 (165.7, 202.6) | 651 (52.5%) |
| 60-69 | 14356 | 1847 | 176 (171.0, 180.6) | 88 (85.5, 90.3) | 1629 (88.2%) | 12517 (87.2%) | 146 (140.8, 149.8) | 80 (77.5, 82.4) | 1564 (84.7%) |
| Total | 50000 | 3573 | 412 (398.4, 426.3) | 206 (199.2, 213.1) | 2426 (67.9%) | 34777 (69.6%) | 271 (260.7, 281.1) | 149 (143.4, 154.6) | 2335 (65.3%) |
| **Women** | | | | | | | | | |
| 40-49 | 18107 | 269 | 13462 (0.0, 19743.6) | 6731 (0.0, 9871.8) | 27 (10.0%) | 3233 (17.9%) | 2185 (0.0, 3192.3) | 1202 (0.0, 1755.7) | 27 (10.0%) |
| 50-59 | 17282 | 577 | 2544 (1729.3, 3115.2) | 1272 (864.6, 1557.6) | 136 (23.6%) | 8329 (48.2%) | 1143 (778.9, 1403.4) | 629 (428.4, 771.9) | 132 (23.0%) |
| 60-69 | 14611 | 962 | 457 (418.9, 488.1) | 229 (209.4, 244.1) | 639 (66.4%) | 10459 (71.6%) | 304 (277.4, 324.1) | 167 (152.6, 178.3) | 626 (65.1%) |
| Total | 50000 | 1808 | 1248 (1151.8, 1336.1) | 624 (575.9, 668.0) | 801 (44.3%) | 22021 (44.0%) | 510 (470.8, 547.0) | 280 (258.9, 300.8) | 785 (43.4%) |

Abbreviations: NNS, number needed to screen; NNI, number needed to invite.

Age structure of hypothetical population extrapolated from Office for National Statistics, England, United Kingdom 2015. Expected events at 10 years based on extrapolation of incidence rates from CPRD, 2014-2019. Age group and sex specific prioritisation thresholds were defined as the level such that the expected false negative rate was controlled to be 5%. NNI and NNS assumes 50% statin compliance. NNI assumes a 50% invitation uptake if assessing without using prioritisation tool, and a 55% invitation uptake if assessing with using prioritisation tool.

**Supplementary table 15:** Number needed to invite and screen to prevent one event, and number of events identified when prioritising with PRS + age, assuming statin compliance of 50%, in a hypothetical population of 100,000 individuals in England.

| | | | **No prioritisation tool used: formal assessment with conventional risk factors + PRS used for all individuals** | | | **Prioritisation using PRS + age, followed by formal assessment with conventional risk factors + PRS** | | | |
|---|---|---|---|---|---|---|---|---|---|
| Age group | Participants | Expected number of events in 10 years | NNI (95% CI) | NNS (95% CI) | Number of events identified as high risk (%) | Participants prioritised (%) | NNI (95% CI) | NNS (95% CI) | Number of events identified as high risk (%) |
| **Men** | | | | | | | | | |
| 40-49 | 18253 | 485 | 2149 (1557.2, 2547.5) | 1074 (778.6, 1273.8) | 170 (35.1%) | 13525 (74.1%) | 1478 (1088.5, 1752.7) | 813 (598.7, 964.0) | 166 (34.3%) |
| 50-59 | 17391 | 1240 | 494 (448.6, 534.4) | 247 (224.3, 267.2) | 705 (56.9%) | 13456 (77.4%) | 364 (328.9, 394.1) | 200 (180.9, 216.7) | 673 (54.3%) |
| 60-69 | 14356 | 1847 | 181 (175.5, 186.7) | 91 (87.7, 93.3) | 1582 (85.7%) | 12424 (86.5%) | 149 (143.3, 153.9) | 82 (78.8, 84.6) | 1517 (82.1%) |
| Total | 50000 | 3573 | 407 (393.0, 422.0) | 204 (196.5, 211.0) | 2457 (68.8%) | 39405 (78.8%) | 304 (292.3, 315.5) | 167 (160.8, 173.5) | 2356 (65.9%) |
| **Women** | | | | | | | | | |
| 40-49 | 18107 | 269 | 11538 (0.0, 16718.6) | 5769 (0.0, 8359.3) | 31 (11.5%) | 11442 (63.2%) | 7733 (0.0, 11331.6) | 4253 (0.0, 6232.4) | 27 (10.0%) |
| 50-59 | 17282 | 577 | 2077 (1455.0, 2472.3) | 1038 (727.5, 1236.2) | 166 (28.8%) | 13590 (78.6%) | 1516 (1050.2, 1820.7) | 834 (577.6, 1001.4) | 163 (28.3%) |
| 60-69 | 14611 | 962 | 452 (418.5, 481.4) | 226 (209.3, 240.7) | 646 (67.2%) | 12357 (84.6%) | 360 (333.3, 383.7) | 198 (183.3, 211.0) | 623 (64.8%) |
| Total | 50000 | 1808 | 1185 (1090.5, 1261.7) | 592 (545.2, 630.8) | 844 (46.7%) | 37389 (74.8%) | 836 (768.7, 893.4) | 460 (422.8, 491.4) | 813 (45.0%) |

Abbreviations: NNS, number needed to screen; NNI, number needed to invite; PRS, polygenic risk score.

Age structure of hypothetical population extrapolated from Office for National Statistics, England, United Kingdom 2015. Expected events at 10 years based on extrapolation of incidence rates from CPRD, 2014-2019. Age group and sex specific prioritisation thresholds were defined as the level such that the expected false negative rate was controlled to be 5%. NNI and NNS assumes 100% statin compliance. NNI assumes a 50% invitation uptake if assessing without using prioritisation tool, and a 55% invitation uptake if assessing with using prioritisation tool.

**Supplementary table 16:** Number needed to invite and screen to prevent one event, and number of events identified when prioritising with PRS and primary care records, assuming statin compliance of 50%, in a hypothetical population of 100,000 individuals in England.

| | | | **No prioritisation tool used: formal assessment with conventional risk factors + PRS used for all individuals** | | | **Prioritisation using PRS and primary care records, followed by formal assessment with conventional risk factors + PRS** | | | |
|---|---|---|---|---|---|---|---|---|---|
| Age group | Participants | Expected number of events in 10 years | NNI (95% CI) | NNS (95% CI) | Number of events identified as high risk (%) | Participants prioritised (%) | NNI (95% CI) | NNS (95% CI) | Number of events identified as high risk (%) |
| **Men** | | | | | | | | | |
| 40-49 | 18253 | 485 | 2149 (1557.2, 2547.5) | 1074 (778.6, 1273.8) | 170 (35.1%) | 7930 (43.4%) | 885 (640.1, 1051.0) | 487 (352.1, 578.0) | 163 (33.6%) |
| 50-59 | 17391 | 1240 | 494 (448.6, 534.4) | 247 (224.3, 267.2) | 705 (56.9%) | 10622 (61.1%) | 288 (259.9, 312.3) | 159 (143.0, 171.8) | 670 (54.0%) |
| 60-69 | 14356 | 1847 | 181 (175.5, 186.7) | 91 (87.7, 93.3) | 1582 (85.7%) | 11436 (79.7%) | 135 (130.4, 139.7) | 75 (71.7, 76.9) | 1535 (83.1%) |
| Total | 50000 | 3573 | 407 (393.0, 422.0) | 204 (196.5, 211.0) | 2457 (68.8%) | 29988 (60.0%) | 230 (221.3, 239.5) | 127 (121.7, 131.7) | 2367 (66.3%) |
| **Women** | | | | | | | | | |
| 40-49 | 18107 | 269 | 11538 (0.0, 16718.6) | 5769 (0.0, 8359.3) | 31 (11.5%) | 3513 (19.4%) | 2035 (0.0, 2953.9) | 1119 (0.0, 1624.7) | 31 (11.7%) |
| 50-59 | 17282 | 577 | 2077 (1455.0, 2472.3) | 1038 (727.5, 1236.2) | 166 (28.8%) | 7616 (44.1%) | 867 (613.9, 1038.0) | 477 (337.6, 570.9) | 160 (27.7%) |
| 60-69 | 14611 | 962 | 452 (418.5, 481.4) | 226 (209.3, 240.7) | 646 (67.2%) | 9872 (67.6%) | 283 (261.8, 301.7) | 156 (144.0, 165.9) | 634 (65.9%) |
| Total | 50000 | 1808 | 1185 (1090.5, 1261.7) | 592 (545.2, 630.8) | 844 (46.7%) | 21001 (42.0%) | 463 (426.0, 495.6) | 255 (234.3, 272.6) | 825 (45.6%) |

Abbreviations: NNS, number needed to screen; NNI, number needed to invite; PRS, polygenic risk score.

Age structure of hypothetical population extrapolated from Office for National Statistics, England, United Kingdom 2015. Expected events at 10 years based on extrapolation of incidence rates from CPRD, 2014-2019. Age group and sex specific prioritisation thresholds were defined as the level such that the expected false negative rate was controlled to be 5%. NNI and NNS assumes 100% statin compliance. NNI assumes a 50% invitation uptake if assessing without using prioritisation tool, and a 55% invitation uptake if assessing with using prioritisation tool.

**Supplementary figure 1**: Flowchart showing selection of patient records for generating summary statistics from CPRD.

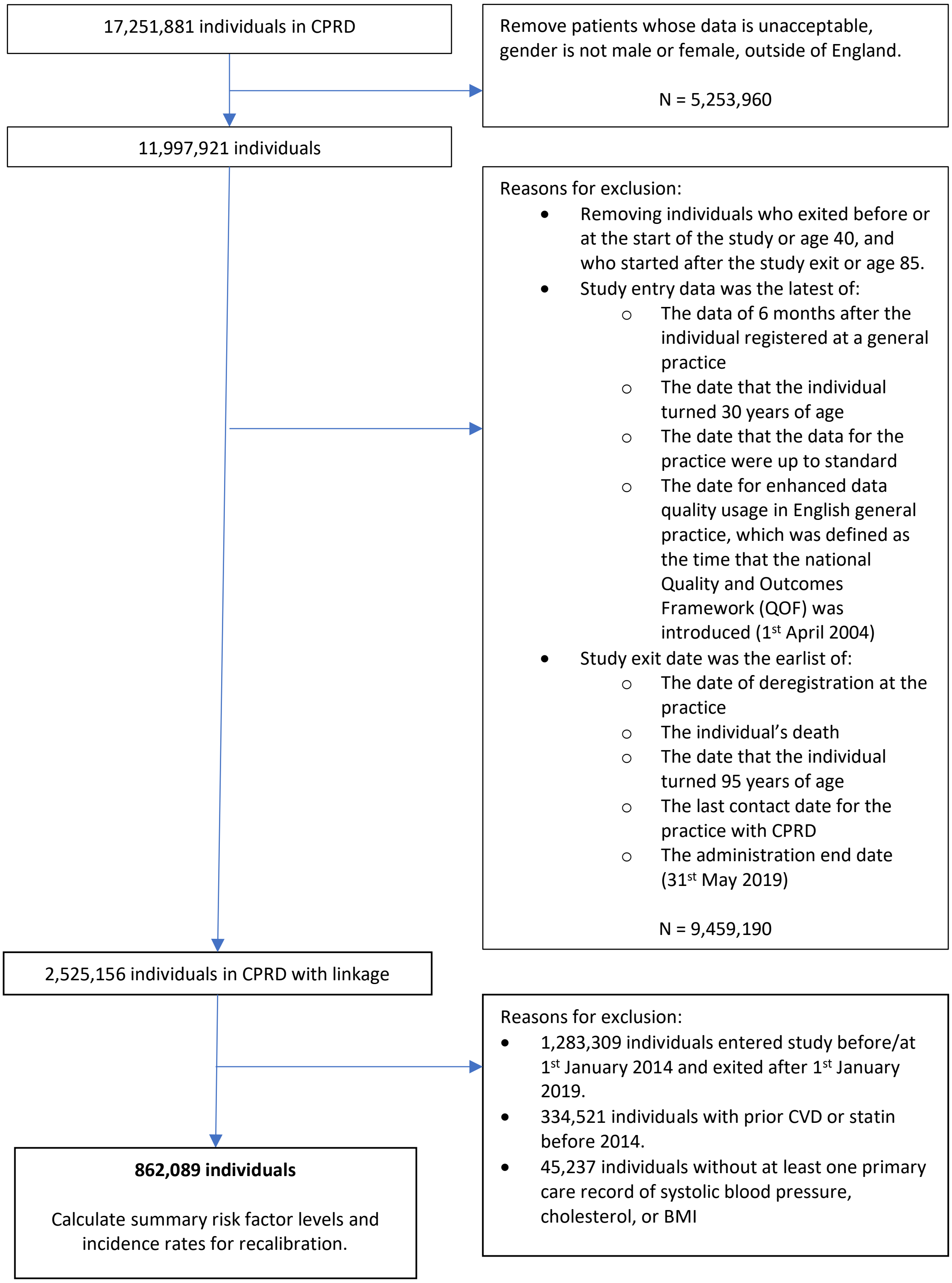

Abbreviations: BMI, body mass index; CVD, cardiovascular disease.

Highlighted in red were individuals without necessary primary care records to calculate incidence rates for sensitivity analyses and were included for sensitivity analysis incidence rates calculation.

**Supplementary figure 2:** Flowchart showing selection of patient records for derivation and population health modelling in UK Biobank.

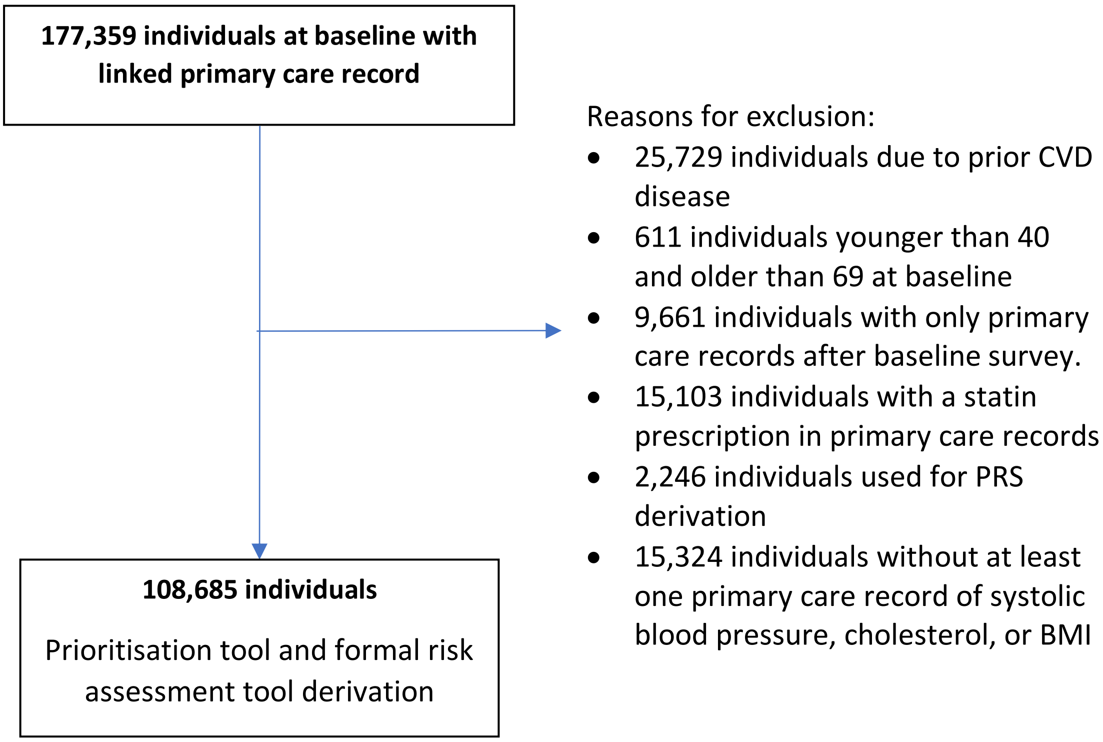

Abbreviations: BMI, body mass index; CVD, cardiovascular disease; PRS, polygenic risk score.

Highlighted in red are individuals without necessary primary care records for primary care based prioritisation tool that were formally assessed in sensitivity analysis.

**Supplementary figure 3:** Age group and sex specific distributions of rescaled 10-year risks for each prioritisation tool and formal assessment tool.

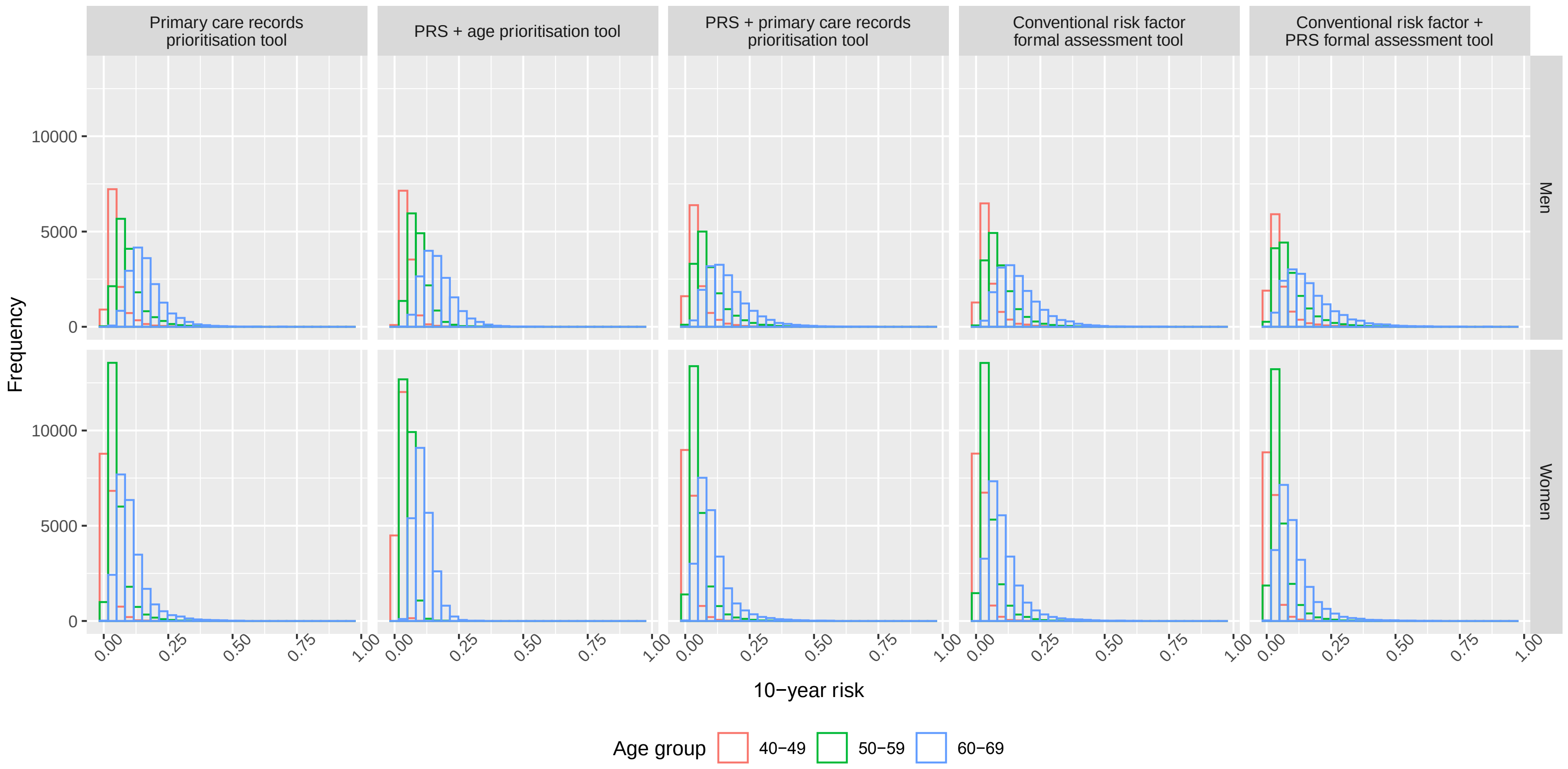

**Supplementary figure 4:** Number needed to invite and screen to prevent one event and number of events identified after prioritisation and formal assessment in a hypothetical population of 100,000 individuals in England, with prioritisation thresholds selected to identify the same number of events if prioritising with primary care records with prioritisation thresholds controlling the false negative rate to 5%.

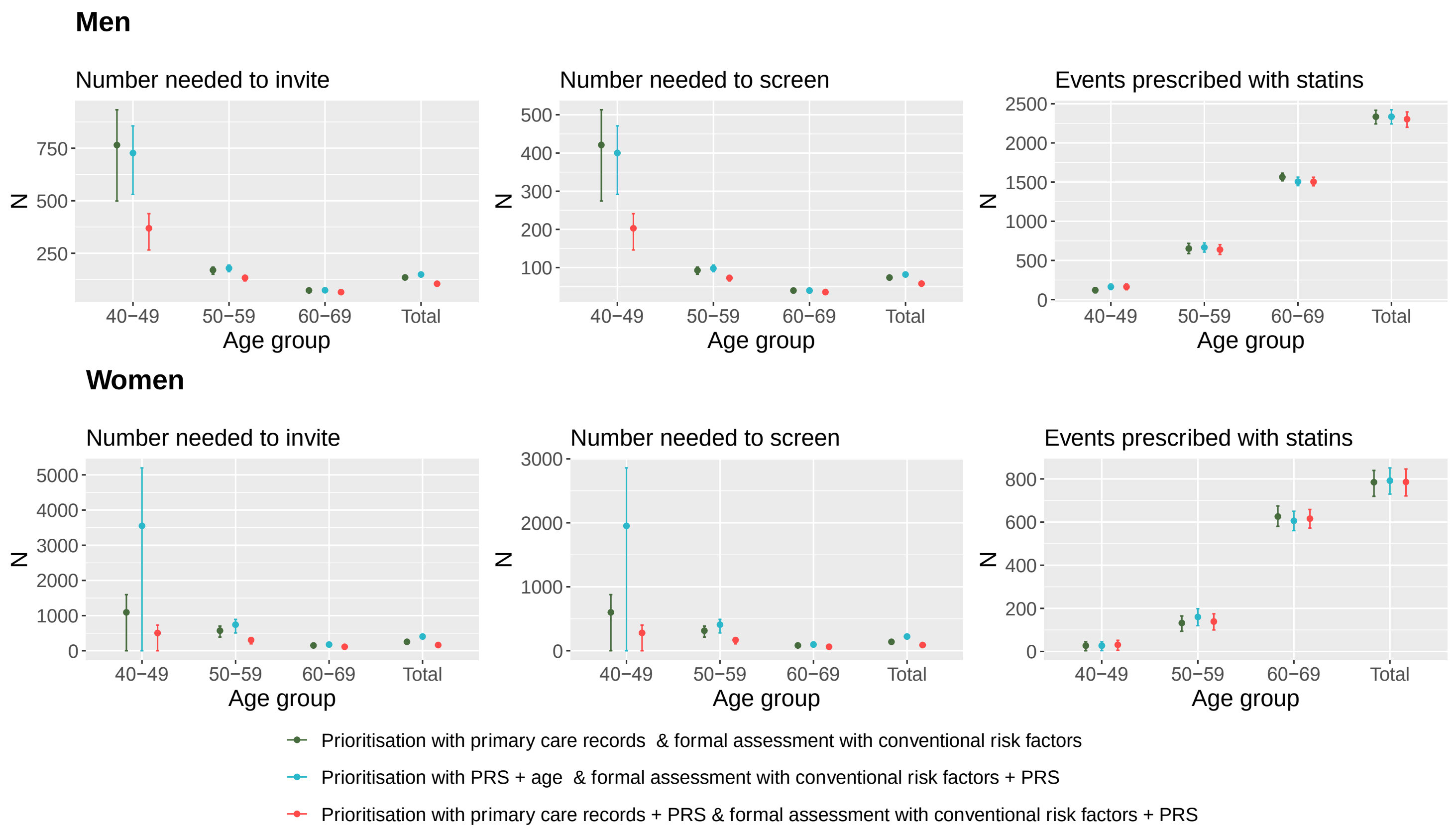

Abbreviations: NNS, number needed to screen; NNI, number needed to invite; PRS, polygenic risk score.

95% confidence intervals are represented by vertical lines. Age group and sex specific prioritisation thresholds were defined as the level such that the expected false negative rate was controlled to be 5%. NNI and NNS assumes 100% statin compliance, and half of all individuals invited for formal assessment attend.

**Supplementary figure 5:** Number needed to invite, number needed to screen and number of events identified after prioritising for a formal CVD assessment, including all individuals without a primary care record for any one of SBP, HDL, total cholesterol or BMI, in a hypothetical population of 100,000 individuals in England.

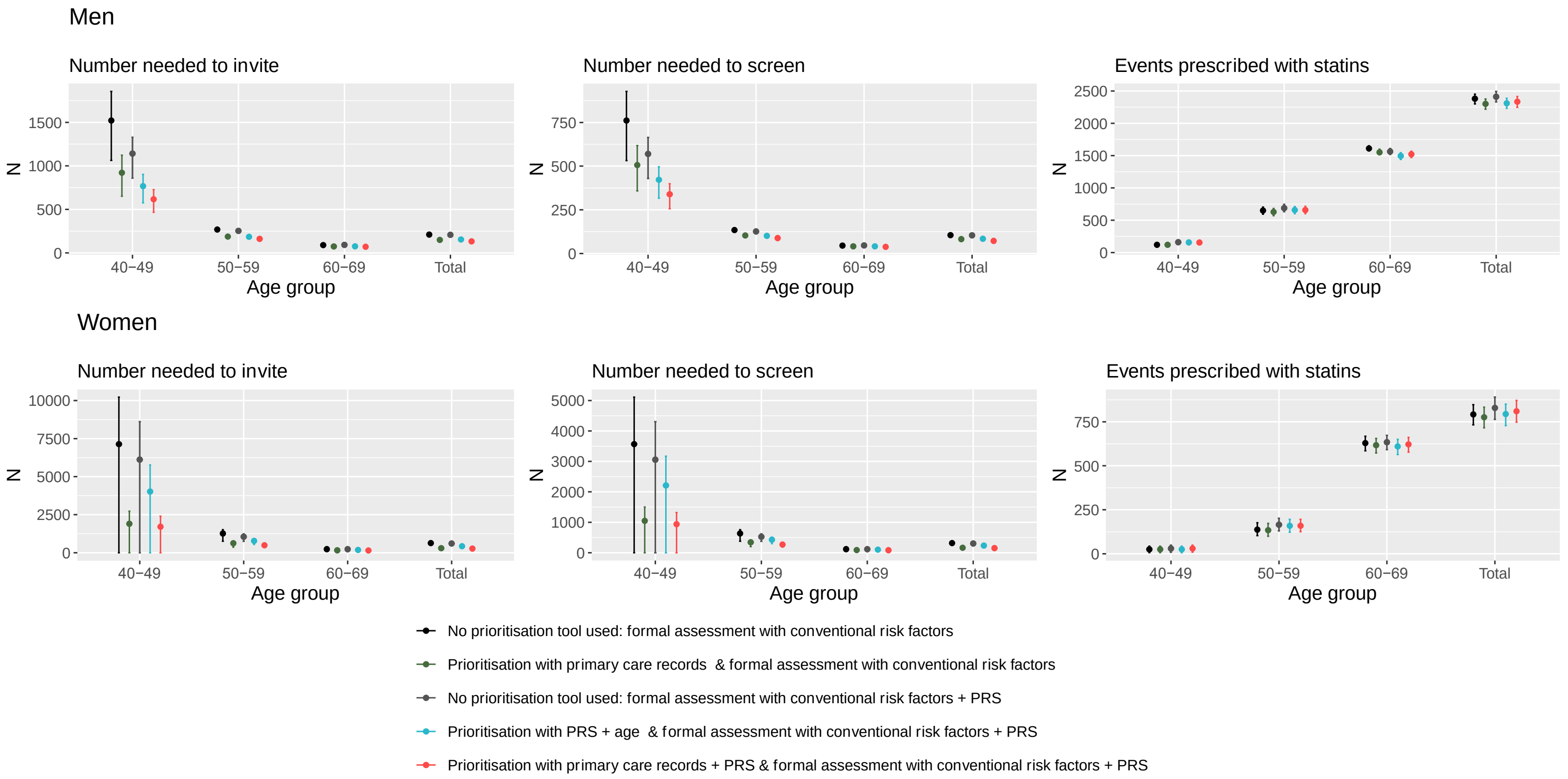

Abbreviations: HDL, high-density lipoprotein; NNI, number needed to invite; NNS, number needed to screen; PRS, polygenic risk score; SBP, systolic blood pressure.

95% confidence intervals are represented by vertical lines. Age group and sex specific prioritisation thresholds were defined as the level such that the expected false negative rate was controlled to be 5%. NNI and NNS assumes 100% statin compliance, and half of all individuals invited for formal assessment attend.

**Supplementary figure 6:** Number needed to invite, number needed to screen and number of events identified after prioritising for a formal CVD assessment, assuming statin compliance of 50%, in a hypothetical population of 100,000 individuals in England.

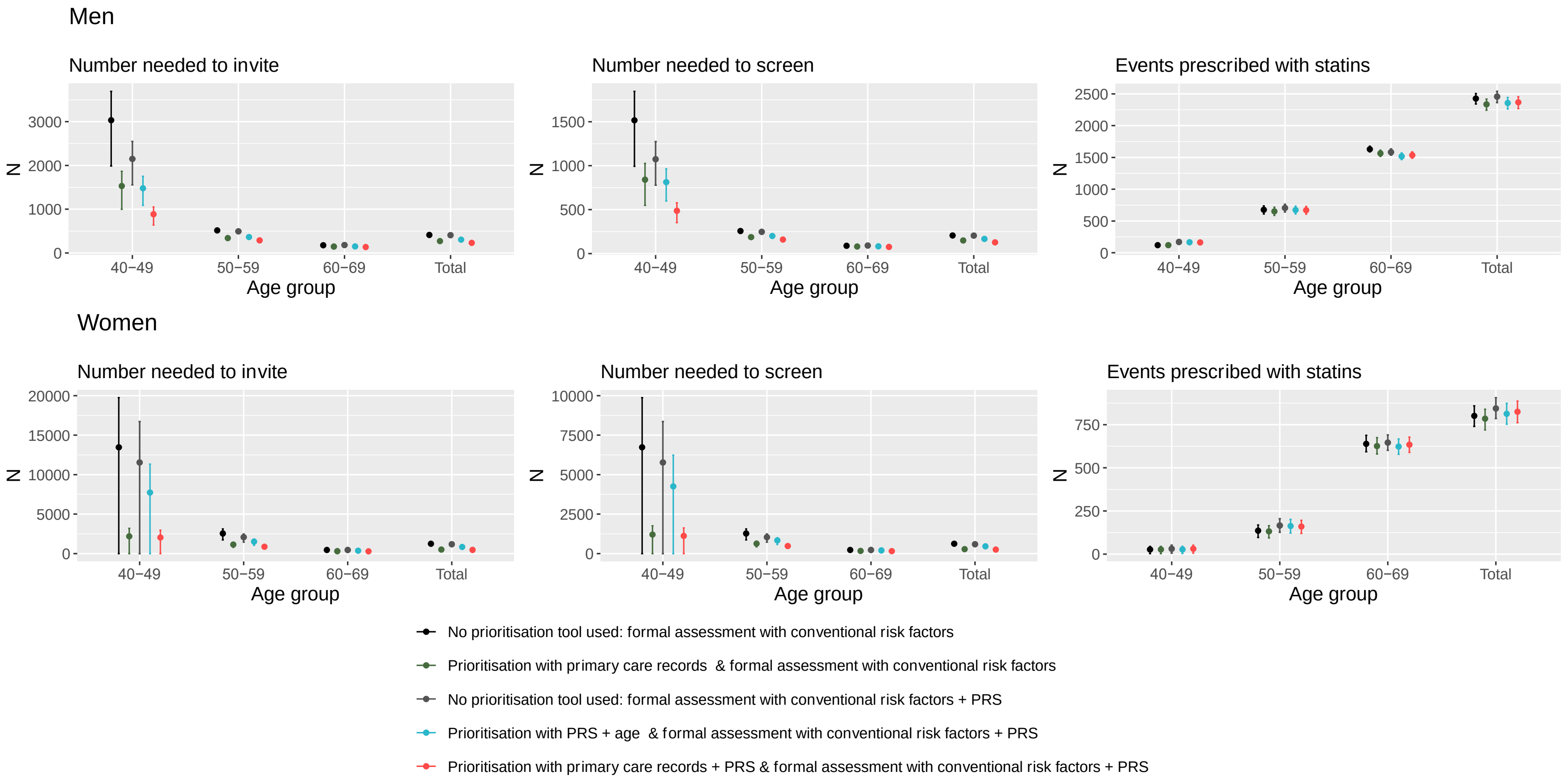

Abbreviations: NNS, number needed to screen; NNI, number needed to invite; PRS, polygenic risk score.

95% confidence intervals are represented by vertical lines. Age group and sex specific prioritisation thresholds were defined as the level such that the expected false negative rate was controlled to be 5%. NNI and NNS assumes 50% statin compliance, and half of all individuals invited for formal assessment attend.

**Web appendix 1.** Summarising repeated measures of risk factors in primary care records using multivariate mixed-effects linear regression models.

**Derivation of prioritisation tools using primary care records.**

Our aim was to derive a prioritisation tool to estimate a 10-year CVD risk for CVD event free individuals using their primary care records only and to then compare with a PRS-based prioritisation tool and one that combines the two together. When utilising longitudinal data in primary care records, the model needs to handle the repeated and sporadic structure of the data.

We used the same risk factors as used in the QRISK2 risk score to derive the primary care records-based prioritisation tool. Values of systolic blood pressure, total and HDL cholesterol, BMI and Townsend score were standardised using sex-specific means and standard deviations. All remaining risk factors were listed as indicator variables and were set to zero until a health record indicated otherwise.

Using the primary care records before baseline survey in a population without prior CVD or diabetes, but including those with prior statin usage, sex-specific multivariate mixed effects models with a fixed slope, random intercept and an intra-correlation structure were used to estimate the risk factor levels at the same timepoint as when the individual attended the UK Biobank baseline assessment. Let $SBP_{ij}$, $Total\ cholesterol_{ij}$, $HDL\ cholesterol_{ij}$, $BMI_{ij}$, $age_{ij}$, $age_{ij}^2$, $AHM_{ij}$, and $statin_{ij}$ denote, respectively, the repeat measures of systolic blood pressure, total cholesterol, HDL cholesterol, BMI, age at visit in years, age at visit in years squared, an indicator for history of anti-hypertensive medication, and an indicator for history of statin medication for individual $i$ and measurement $j$. The sex-specific multivariate mixed models and its corresponding correlated covariance structure were of the following form:

$$SBP_{ij} = a_1 + b_1 age_{ij} + c_1 age_{ij}^2 + (d * AHM_{ij}) + u_{1i} + e_{1ij}$$

$$Total\ cholesterol_{ij} = a_2 + b_2 age_{ij} + c_2 age_{ij}^2 + (e * statin_{ij}) + u_{2i} + e_{2ij}$$

$$HDL\ cholesterol_{ij} = a_3 + b_3 age_{ij} + c_3 age_{ij}^2 + u_{3i} + e_{3ij}$$

$$BMI_{ij} = a_4 + b_4 age_{ij} + c_4 age_{ij}^2 + u_{4i} + e_{4ij}$$

Where $\begin{bmatrix} u_{1i} \\ u_{2i} \\ u_{3i} \\ u_{4i} \end{bmatrix} \sim multivariate\ normal \left( \begin{bmatrix} 0 \\ 0 \\ 0 \\ 0 \end{bmatrix}, \begin{bmatrix} \sigma_1^2 & \sigma_{12} & \sigma_{13} & \sigma_{14} \\ \sigma_{12} & \sigma_2^2 & \sigma_{23} & \sigma_{24} \\ \sigma_{13} & \sigma_{23} & \sigma_3^2 & \sigma_{34} \\ \sigma_{14} & \sigma_{24} & \sigma_{34} & \sigma_4^2 \end{bmatrix} \right)$

And $\begin{bmatrix} e_{1ij} \\ e_{2ij} \\ e_{3ij} \\ e_{4ij} \end{bmatrix} \sim multivariate\ normal \left( \begin{bmatrix} 0 \\ 0 \\ 0 \\ 0 \end{bmatrix}, \begin{bmatrix} \sigma_{e1}^2 & 0 & 0 & 0 \\ 0 & \sigma_{e2}^2 & 0 & 0 \\ 0 & 0 & \sigma_{e3}^2 & 0 \\ 0 & 0 & 0 & \sigma_{e4}^2 \end{bmatrix} \right)$

Where $u_{1i}$ to $u_{4i}$ represent the random intercepts but are correlated between risk factors. $e_{1ij}$ to $e_{4ij}$ represents the uncorrelated residual errors for each risk factor.

A mixed effects model was chosen to take into account the sporadic nature of electronic health records, as well as being able to model the intra-correlations between each risk factor. In addition, the model only needs a minimum of one recorded measurement of any one risk factor to estimate all four of the risk factors.

The model assumes that all risk factors jointly follow a multivariate normal distribution. Inference based from the multivariate normal distribution may often be reasonable even if the multivariate normality does not hold, especially in the context of imputation of missing data[1] and regression calibration[2,3].

**Web appendix 2.** Rescaling of prioritisation tool and formal risk assessment tool risks for population health modelling.

Our aim was to validate each prioritisation tool to estimate the health impact in a general population in England. We used UK Biobank due to its availability of detailed measurements at baseline, which was used to estimate a 10-year formal assessment risk, but also genetic data and linked historical primary care records necessary for the formal assessment model using conventional risk factors and PRS, and the prioritisation tools derived using primary care records and/or PRS. The breadth of the data allowed for a direct comparison of each prioritisation tool in the same individuals.

However, UKB participants have been shown to be healthier than the general population both in terms of risk factor levels and CVD incidence rates. Deriving and modelling the health impact of all prioritisation tools and formal risk tools in UK Biobank without adjustments would lead to a biased distribution of 10-year risks estimated, with the distribution of risks being skewed to the right and be narrow relative to the distribution observed in the general population. To more accurately use UK Biobank for population health modelling, the distribution of 10-year risks estimated were rescaled.

Rescaling was completed for each tool and by sex, using methods similar to those previously described[4], and allowed the mean level of predicted risks based on UKB data to match what was observed in CPRD. We used sex-specific mean risk factor levels calculated from the Clinical Practice Research Datalink (CPRD) between the years 2014 and 2019 within 5-year age groups to estimate the predicted risk in the general population by fitting the average level risk factors into the published QRISK2 risk model **(supplementary table 2).** This allows us to calculate scaling factors to rescale each prioritisation tool and formal risk assessment model to have a distribution similar to what would be expected in the general population.

A linear model was fit within each tool and by sex to relate the observed risk ($\theta_{obs}$) and predicted risk ($\theta_{pred}$) estimated for each 5-year age group ($c_s$):

$$\log_e(-\log_e(1-\theta_{obs,c_s})) = \beta_0 + \beta_1 \times \log_e(-\log_e(1-\theta_{pred,c_s}))$$

The estimated $\beta_0$ and $\beta_1$ were then used as scaling factors to rescale each individual's original 10-year risk ($\theta_{pred,i}$) to give a new rescaled estimate $\theta_{newpred,i}$ :

$$\theta_{newpred,i} = 1 - \exp(-\exp(\beta_0 + \beta_1 \times \log_e(-\log_e(1-\theta_{pred,i}))))$$

**Web Appendix 3.** The RECORD statement[5]– checklist of items, extended from the STROBE statement, that should be reported in observational studies using routinely collected health data.

| | Item No. | STROBE items | Location in manuscript where items are reported | RECORD items | Location in manuscript where items are reported |
|---|---|---|---|---|---|
| **Title and abstract** | | | | | |
| | 1 | (a) Indicate the study's design with a commonly used term in the title or the abstract (b) Provide in the abstract an informative and balanced summary of what was done and what was found | 3 | RECORD 1.1: The type of data used should be specified in the title or abstract. When possible, the name of the databases used should be included.<br><br>RECORD 1.2: If applicable, the geographic region and timeframe within which the study took place should be reported in the title or abstract.<br><br>RECORD 1.3: If linkage between databases was conducted for the study, this should be clearly stated in the title or abstract. | 3<br><br>3<br><br>3 |
| **Introduction** | | | | | |
| Background rationale | 2 | Explain the scientific background and rationale for the | 4 | | |

| | | | | | |
|---|---|---|---|---|---|
| | | investigation being reported | | | |
| Objectives | 3 | State specific objectives, including any prespecified hypotheses | 4 | | |
| **Methods** | | | | | |
| Study Design | 4 | Present key elements of study design early in the paper | 5 | | |
| Setting | 5 | Describe the setting, locations, and relevant dates, including periods of recruitment, exposure, follow-up, and data collection | 5 | | |
| Participants | 6 | *(a) Cohort study* - Give the eligibility criteria, and the sources and methods of selection of participants. Describe methods of follow-up *Case-control study* - Give the eligibility criteria, and the sources and methods of case ascertainment and control selection. Give the rationale for the choice of cases and controls | 5 | RECORD 6.1: The methods of study population selection (such as codes or algorithms used to identify subjects) should be listed in detail. If this is not possible, an explanation should be provided.<br><br>RECORD 6.2: Any validation studies of the codes or algorithms used to select the population should be | 6<br><br>6 |

| | | | | | |
|---|---|---|---|---|---|
| | | *Cross-sectional study* - Give the eligibility criteria, and the sources and methods of selection of participants<br><br>*(b) Cohort study* - For matched studies, give matching criteria and number of exposed and unexposed<br>*Case-control study* - For matched studies, give matching criteria and the number of controls per case | | referenced. If validation was conducted for this study and not published elsewhere, detailed methods and results should be provided.<br><br>RECORD 6.3: If the study involved linkage of databases, consider use of a flow diagram or other graphical display to demonstrate the data linkage process, including the number of individuals with linked data at each stage. | <br><br><br><br><br><br><br><br><br><br><br>Supplementary figure 2 |
| Variables | 7 | Clearly define all outcomes, exposures, predictors, potential confounders, and effect modifiers. Give diagnostic criteria, if applicable. | 6 | RECORD 7.1: A complete list of codes and algorithms used to classify exposures, outcomes, confounders, and effect modifiers should be provided. If these cannot be reported, an explanation should be provided. | 6, supplementary table 1 |
| Data sources/ measurement | 8 | For each variable of | 6 | | |

| | | | |
|---|---|---|---|
| | | interest, give sources of data and details of methods of assessment (measurement). Describe comparability of assessment methods if there is more than one group | |
| Bias | 9 | Describe any efforts to address potential sources of bias | 7 |
| Study size | 10 | Explain how the study size was arrived at | Supplementary figures 1-2 |
| Quantitative variables | 11 | Explain how quantitative variables were handled in the analyses. If applicable, describe which groupings were chosen, and why | 6-7 |
| Statistical methods | 12 | (a) Describe all statistical methods, including those used to control for confounding<br>(b) Describe any methods used to examine subgroups and interactions<br>(c) Explain how missing data were addressed<br>(d) *Cohort study* - If applicable, explain how loss | 6-8 |

| | | | | | |
|---|---|---|---|---|---|
| | | to follow-up was addressed<br>*Case-control study* - If applicable, explain how matching of cases and controls was addressed<br>*Cross-sectional study* - If applicable, describe analytical methods taking account of sampling strategy<br>(e) Describe any sensitivity analyses | | | |
| Data access and cleaning methods | | N/A | | RECORD 12.1: Authors should describe the extent to which the investigators had access to the database population used to create the study population.<br><br>RECORD 12.2: Authors should provide information on the data cleaning methods used in the study. | 5<br><br>5, Supplementary figures 1-2 |
| Linkage | | N/A | | RECORD 12.3: State whether the study included | 6 |

| | | | | | |
|---|---|---|---|---|---|
| | | | | person-level, institutional-level, or other data linkage across two or more databases. The methods of linkage and methods of linkage quality evaluation should be provided. | |
| **Results** | | | | | |
| Participants | 13 | (a) Report the numbers of individuals at each stage of the study (*e.g.*, numbers potentially eligible, examined for eligibility, confirmed eligible, included in the study, completing follow-up, and analysed)<br>(b) Give reasons for non-participation at each stage.<br>(c) Consider use of a flow diagram | 9, Supplementary figures 1-2 | RECORD 13.1: Describe in detail the selection of the persons included in the study (*i.e.,* study population selection) including filtering based on data quality, data availability and linkage. The selection of included persons can be described in the text and/or by means of the study flow diagram. | 5, Supplementary figures 1-2 |
| Descriptive data | 14 | (a) Give characteristics of study participants (*e.g.*, demographic, clinical, social) and information on exposures | Table 1 | | |

| | | | |
|---|---|---|---|
| | | and potential confounders<br>(b) Indicate the number of participants with missing data for each variable of interest<br>(c) *Cohort study* - summarise follow-up time (*e.g.*, average and total amount) | |
| Outcome data | 15 | *Cohort study* - Report numbers of outcome events or summary measures over time<br>*Case-control study* - Report numbers in each exposure category, or summary measures of exposure<br>*Cross-sectional study* - Report numbers of outcome events or summary measures | 9 |
| Main results | 16 | (a) Give unadjusted estimates and, if applicable, confounder-adjusted estimates and their precision (e.g., 95% confidence interval). Make clear which | Supplementary table 4 |

| | | | | | |
|---|---|---|---|---|---|
| | | confounders were adjusted for and why they were included<br>(b) Report category boundaries when continuous variables were categorized<br>(c) If relevant, consider translating estimates of relative risk into absolute risk for a meaningful time period | | | |
| Other analyses | 17 | Report other analyses done—e.g., analyses of subgroups and interactions, and sensitivity analyses | 10-12, Table 4, Supplementary figures 5-6 | | |
| **Discussion** | | | | | |
| Key results | 18 | Summarise key results with reference to study objectives | 13 | | |
| Limitations | 19 | Discuss limitations of the study, taking into account sources of potential bias or imprecision. Discuss both direction and magnitude of any potential bias | 14 | RECORD 19.1: Discuss the implications of using data that were not created or collected to answer the specific research question(s). Include discussion of misclassification bias, | 14 |

| | | | | | |
|---|---|---|---|---|---|
| | | | | unmeasured confounding, missing data, and changing eligibility over time, as they pertain to the study being reported. | |
| Interpretation | 20 | Give a cautious overall interpretation of results considering objectives, limitations, multiplicity of analyses, results from similar studies, and other relevant evidence | 13 | | |
| Generalisability | 21 | Discuss the generalisability (external validity) of the study results | 13-14 | | |
| **Other Information** | | | | | |
| Funding | 22 | Give the source of funding and the role of the funders for the present study and, if applicable, for the original study on which the present article is based | 16 | | |
| Accessibility of protocol, raw data, and programming code | | N/A | | RECORD 22.1: Authors should provide information on how to access any supplemental information | 17 |

| | | | | |
|---|---|---|---|---|
| | | | | such as the study protocol, raw data, or programming code. |
